# EquiOx: A Prospective study of pulse oximeter bias and skin pigmentation in critically-ill adults

**DOI:** 10.1101/2025.10.06.25337217

**Authors:** Carolyn M. Hendrickson, Michael S. Lipnick, David Chen, Danni Chen, Tyler J. Law, Romain Pirracchio, John R. Feiner, Ella Behnke, Yu (Celine) Chou, Seif Elmankabadi, Caroline Hughes, Kelvin L. Moore, Lily Ortiz, Leonid Shmuylovich, Malvina B. Eydelman, Gregory Leeb, Ellis P. Monk, Olubunmi Okunlola, Daryl Dorsey, Jaaie Varshney, Fekir Negussie, Philip E. Bickler

## Abstract

**Rationale:** Pulse oximeter performance may vary by skin pigmentation, but most data are retrospective with key limitations.

**Objective:** To quantify pulse oximeter bias [mean difference between pulse oximeter oxygen saturation (SpO_2_) and arterial blood functional oxygen saturation (SaO_2_)], and average root mean square error (A_RMS_) and estimate adjusted effects of skin pigment on bias or A_RMS_ in critically-ill adults

**Methods:** Prospective single-center study of 631 ICU patients (2022–2024) directly observed SpO and SaO pairs. Skin pigment was assessed using the subjective Monk Skin Tone Scale and objective spectrophotometry measurement (Individual Typology Angle [ITA]). Adjusted effects were estimated with targeted maximum likelihood estimation.

**Main Results:** Among 1,760 paired measurements from 631 critically-ill adults, median SaO_2_ was 98% (IQR 96%, 99%) with 40 episodes of stable hypoxemia (SpO_2_<90%). SpO_2_ systematically underestimated SaO_2_ [median bias= −1.70 IQR (−2.84, −0.50)]. Bias was less negative in patients with darker skin (ITA <-30°) [−1.05 (−2.44, −0.10)] vs lighter skin (ITA >30°) [−2.01 (−3.34, −1.00)] and remained significantly different in adjusted analyses. A_RMS_ was 3.87 (95% CI 3.25, 4.53) overall and 4.49 (95% CI 2.63, 7.07) in patients with darker skin. Ear probes performed worse than finger probes [bias: −2.20 vs. −1.60; A_RMS_: 4.90 vs. 2.70].

**Conclusions:** In this large ICU cohort, hypoxemia was rare, pulse oximeters systematically underestimated SaO, performance varied by skin pigment and probe site. Bias was less negative in patients with darker skin. Pulse oximeter inaccuracies in ICU patients may be more substantial than clinicians recognize.

**Key Points:** *Question:* Does pulse oximeter bias in critically-ill adults differ by skin pigmentation?

*Findings:* In this prospective, single-center study of 631 critically-ill adults, oximeter mean bias was negative for all patients but relatively less negative for patients with darker pigment. Bias variation by pigment was nonlinear and larger in ear than finger probes.

*Meaning:* Pulse oximeter bias varies with skin pigmentation and may not always be positive in patients with darker pigmentation. Pulse oximeter inaccuracy may be larger than clinicians appreciate. Additional studies with multiple oximeter brands and more stable hypoxemia are needed to further understand bias variation with skin pigment.

## Background

Disparities in pulse oximeter performance for people with darker skin pigment or with Black/African American racial identity have been recognized for decades,^1–3^ but were only recently linked to health outcomes during the COVID-19 pandemic.^4–8^ Pulse oximeters accuracy can differ between real-world patients and healthy volunteers used for regulatory verification testing .^9,10^ Given a vast number of daily clinical decisions that rely on pulse oximeter oxygen saturation (SpO_2_) without arterial blood gas (ABG) functional oxygen saturation (SaO_2_), these performance differences require further investigation.^11^ Most retrospective studies have major limitations, including unadjusted analyses, delays between paired SaO2 and SpO2 readings, conflating race with skin pigmentation, failing to distinguish functional and fractional hemoglobin saturation, and failing to distinguish between probe types.^12–16^

This prospective study (The EquiOx Study - NCT05554510) was designed to test for differences in pulse oximeter bias (SpO_2_-SaO_2_) across varying skin pigmentation among critically-ill adults.

## Methods

### Study Design

This Institutional Review Board (IRB) approved prospective observational cohort study of directly observed paired measurements of arterial blood gas SaO_2_ and pulse oximeter SpO_2_ enrolled adult patients in a mixed medical-surgical intensive care unit (ICU) at a university-affiliated, Level 1 trauma center and safety-net hospital between August 2022 and September 2024. All eligible subjects were approached for participation while study staff were present. A modified approach to informed consent for this minimal risk study was granted by the IRB with an initial waiver when informed consent was not feasible. Informed consent was then obtained from patients or surrogates. The study protocol and statistical analysis plan are available in the Online Data Supplement.

### Study Population

Adults admitted to the ICU with an arterial line for clinical purposes were eligible. Participants were followed for up to two weeks.

### Skin Pigment Assessment

Trained research staff measured skin pigment (eMethods, Online Data Supplement). Objective skin pigment data were measured in triplicate with the Konica Minolta CM-700d spectrophotometer. Analyses used the median of the three measurements of Individual Typology Angle (ITA), a surrogate for melanin content at a site close to the oximeter.^17^ We used previously established ITA cutoffs: dark = ITA <-30°, medium = ITA −30° to 30°, light = ITA >30°).^18,19^ After six months and 120 enrollments, we added subjective skin assessment to the protocol using the Monk Skin Tone (MST) (dark = HIJ, medium = EFG, and light = ABCD).^20–22^

### Data Collection

Research coordinators at the patient’s bedside recorded SpO_2_ and other clinical data including pulse oximeter probe location, participant movement, duration of SpO_2_ stability prior to sampling (defined as SpO_2_ +1% for >30 seconds), amplitude and regularity of plethysmography, blood pressure, perfusion index, vasopressor dose, and oxygen delivery.

Functional arterial oxygen saturation (SaO2) was measured with a GEM 5000 Blood Gas Analyzer (Werfen, Bedford MA). Standard of care patient monitors and probes were placed by the clinical team (eMethods, Online Data Supplement): Philips IntelliVue MX800 monitor (Cambridge, MA) using Masimo SET module technology (Irvine, CA) or standalone monitors for Masimo Radical-7, Radical-57, or Radical-87 (Irvine, CA). Ear probe oximeters were only applied if a finger probe was not consistently reporting SpO2.

Comorbidities and self-identified race and ethnicity were collected with chart review (Epic MyChart, Verona, WI) .^23–26^ All data were entered into a Research Electronic Data Capture (REDCap) Database hosted at University of California San Francisco.^27,28^

### Statistical Analysis

#### Sample Size Calculation

The initial sample size calculations and planned interim revisions by the Data Monitoring Committee (DMC) are detailed in the supplementary materials. A final sample size target of 300-350 participants was recommended. Data collection continued beyond this target until the end of the funding period.

#### Clinical and Demographic Description of Study Population

Data are reported as median (IQR) or n (%) and compared using the Kruskal-Wallis or Chi-square test as appropriate. We used Fisher’s exact test or Monte Carlo simulation when expected counts were <5. We used a likelihood ratio test to compare linear mixed effects models with and without comparison groups for measurement-level covariates. Generalized estimating equations were applied for categorical outcomes. A two-sided P value < 0.05 was considered statistically significant.

#### Pulse oximeter performance

Pulse oximeter bias is SpO_2_ - SaO_2_. Average root mean squared error (A_RMS_), was calculated as the square root of the average of the squared difference between SpO_2_ and SaO_2_. We referenced 2013 FDA recommended thresholds for controlled desaturation studies among healthy volunteers (A_RMS_ ≤3% for finger, ≤3.5% for ear) and 2025 FDA draft guidance (A_RMS_ 95% CI upper limit <3%).^29^ We evaluated sensitivity and specificity for hypoxemia (SaO_2_ <90%). Bland Altman plots were adjusted for repeated measures (Figure E1).^30^ In post-hoc analyses, “occult hypoxemia” was identified as SaO_2_ <88% when SpO_2_ is 92-96%.^12^

#### Association with Skin Pigmentation or Race/Ethnicity

We calculated the observed median pulse oximeter error and A_RMS_ in skin pigment strata and race/ethnicity categories. Using targeted maximum likelihood estimation (TMLE) and the SuperLearner algorithm (eMethods), we estimated the adjusted average effect on mean SpO_2_ bias or A_RMS_ attributable to race/ethnicity or skin pigment categories (ITA or MST).^31,32^ Skin pigment category effects were averaged over the joint covariate distribution of confounders and outcome predictors identified by directed acyclic graphs (DAGs) (Supplement). In our DAG, measurement-level variables (e.g. ear vs finger probe, perfusion index etc.) were assumed to be unaffected by pigment category or race/ethnicity and so are treated as potential confounders rather than as potential mediators in our analyses. Models were adjusted for covariates including age, body mass index, perfusion index, shock present at enrollment, administration of vasopressors, and variety of cardiovascular diseases (Table E1). For the models, measurement-level variables (e.g. ear vs finger probe, perfusion index etc.) were assumed to be unaffected by pigment category or race/ethnicity. Estimated differences between groups along the pigmentation scales were used to test for differential bias while controlling family-wise error rate with the Holm-Bonferroni method where q-values < 0.05 were considered statistically significant. In post hoc analyses we tested for linearity in the relationship between ITA and bias using the Harvey-Collier and rainbow tests.

### Missing Data

Missing data was imputed with predictive mean matching for a small proportion of seven covariates (S5, eMethods) but not outcomes or exposures.

### Subgroup Analyses

A “manufacturer guidance” subgroup analysis was planned *a priori* and excluded data that would likely be viewed as unreliable by a clinician at the bedside, such as observations with patient movement, unstable SpO_2_ readings, or nail polish (eMethods). The other three subgroup analyses (SpO_2_ <99%, finger and ear probe observations) were *post hoc*. Figure 1, S5 eMethods)

### Sensitivity Analyses

Sensitivity analyses assessed the robustness of our findings to violations of our causal assumptions (eMethods, Online Data Supplement, including tests for unmeasured confounding). Given a lack of scientific consensus on how to optimally define dark skin pigment by ITA,^22^ we assessed the impact of changing ITA cutoff for “dark” from −30°to - 50° (darker) or −10° (lighter).

Statistical analyses were performed in R (4.3.3) and relied on the tidyverse (2.0.0) and SuperLearner (2.0-29) packages (eMethods, Online Data Supplement).

## Results

### Study population

We enrolled 631 participants who contributed 1,760 paired SaO and SpO measurements with a median of 2 samples (IQR 1–3) across 2.4 days (IQR 0.9–6.5) (Figure 1), with 250 (40%) participants having only 1 sample taken. In the full cohort, 57% had shock at enrollment, 76% of ABGs were taken during mechanical ventilation and 58% were collected on vasopressors (Table 1). Median SaO_2_ was 98% (IQR 96%, 99%), and median SpO_2_ was 96% (IQR 94%, 98%). We observed 40 (2%) hypoxemic episodes (SaO_2_ <90%) and 83 (5%) episodes of stable pulse oximeter desaturation (SpO_2_ <90%). We observed two instances of “occult hypoxemia” (SaO_2_ <88% when SpO_2_ was 92-96%) from two different patients with ear probes (Figure 2). Descriptive statistics of skin pigment and race/ethnicity are presented in Table E3.

**Table 1.**
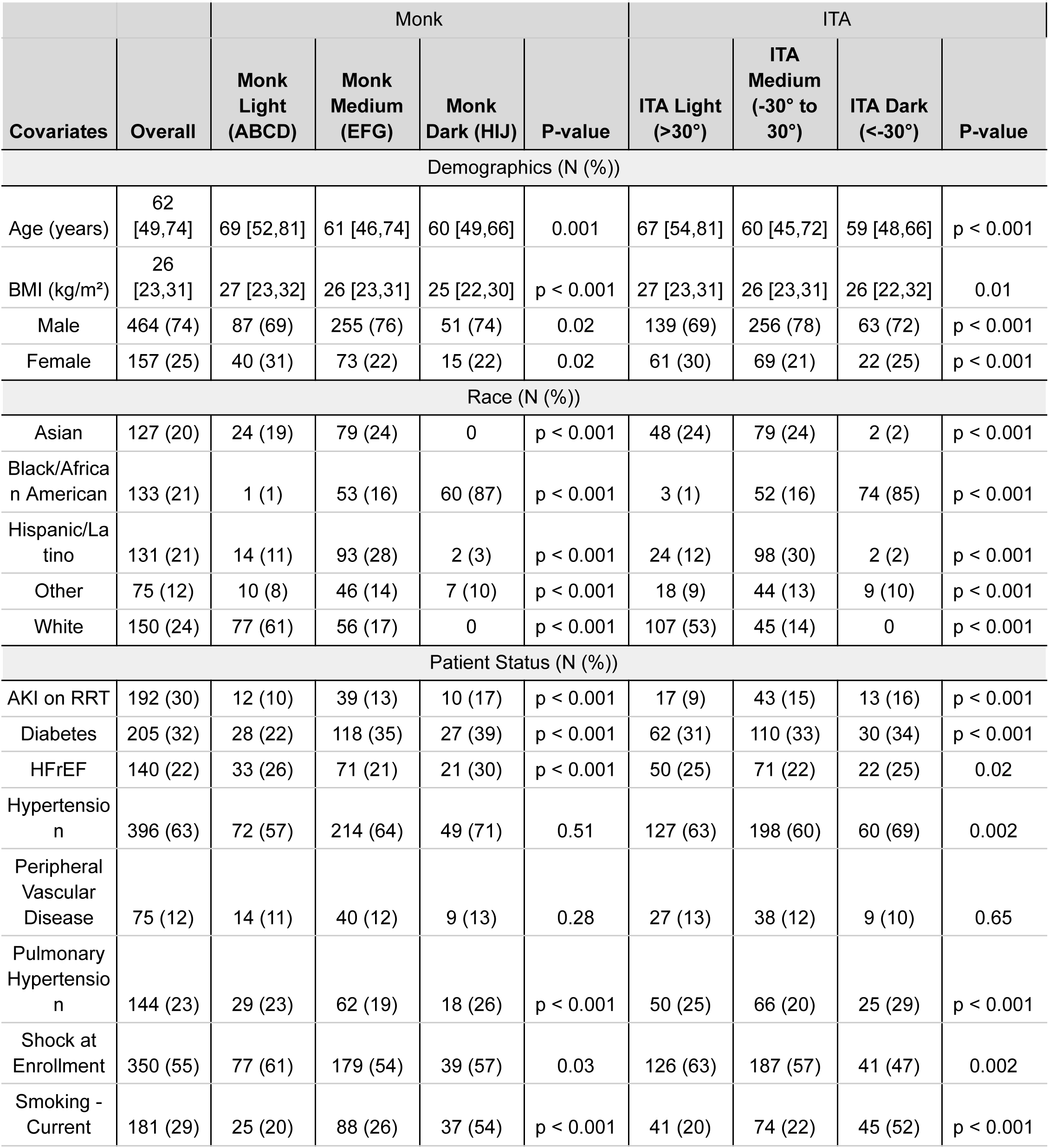
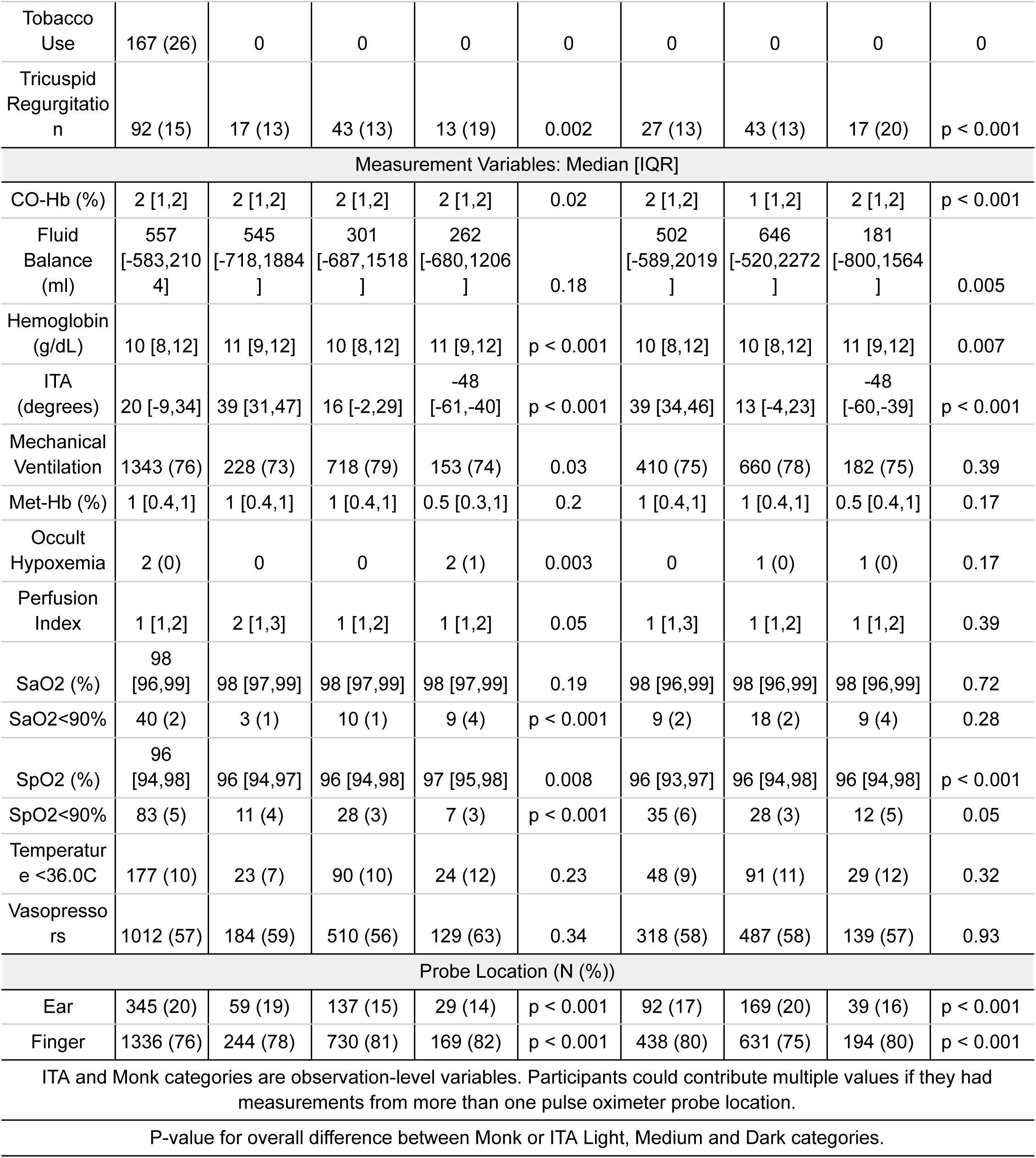
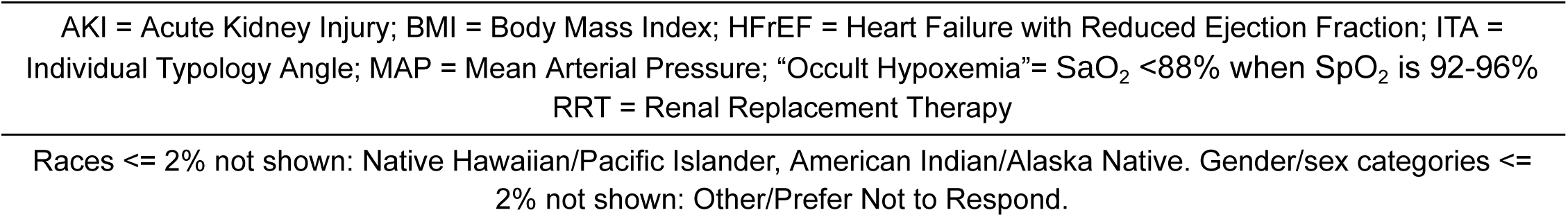
Clinical and Demographic Characteristics of Full Cohort.

### Oximeter Performance

In the full cohort, observed median pulse oximeter bias was negative [median −1.70, IQR (−2.84, −0.50)], meaning SpO2 tended to under-estimate SaO_2_. The A_RMS_ in the full cohort was 3.87 (95% CI 3.25, 4.53). The sensitivity of the pulse oximeters for detecting hypoxemia was 0.72 (95% CI: 0.55, 0.88), and the specificity was 0.97 (95% CI: 0.96, 0.98). In the full cohort, a modified Bland Altman analysis adjusted for repeated measures calculated the limits of agreement (LOA: −6.91%, 2.16%), (Figure E1).

The pulse oximeter bias was positive (over-estimating SaO_2_) in 20% of observations in the full cohort (Figure E2) with 71% of these positive observations in ’manufacturer guidance’ cohort-selected for higher quality data. Characteristics of readings with positive bias vs. non-positive bias (SpO_2_ - SaO_2_ ≤ 0) differ in many respects (Table E4).

The positive bias observations had a higher proportion of Black/African American patients, patients with dark skin, diabetes, hypertension, peripheral vascular disease, and shock at enrollment. Observations with positive bias had lower perfusion index and higher prevalence of hypoxemia (SaO_2_ <90%). Among observations with positive bias, 17 (5%) had hypoxemia (SaO2 <90%). Eight (2%) of positive bias observations had SpO2 >90% but SaO2 <90%.

The median observed oximeter bias was negative for all skin pigment categories in the full cohort and subgroup analyses. While most IQRs of bias were negative, the IQR in the dark MST patients in all groups and in the dark ITA patients with ear probes included positive values (Table 2, Figure E3, Tables E6 - E9). The proportion of positive bias observations was smallest in the light pigment categories and largest in the dark pigment categories (MST and ITA) (Table E4).

**Table 2.**
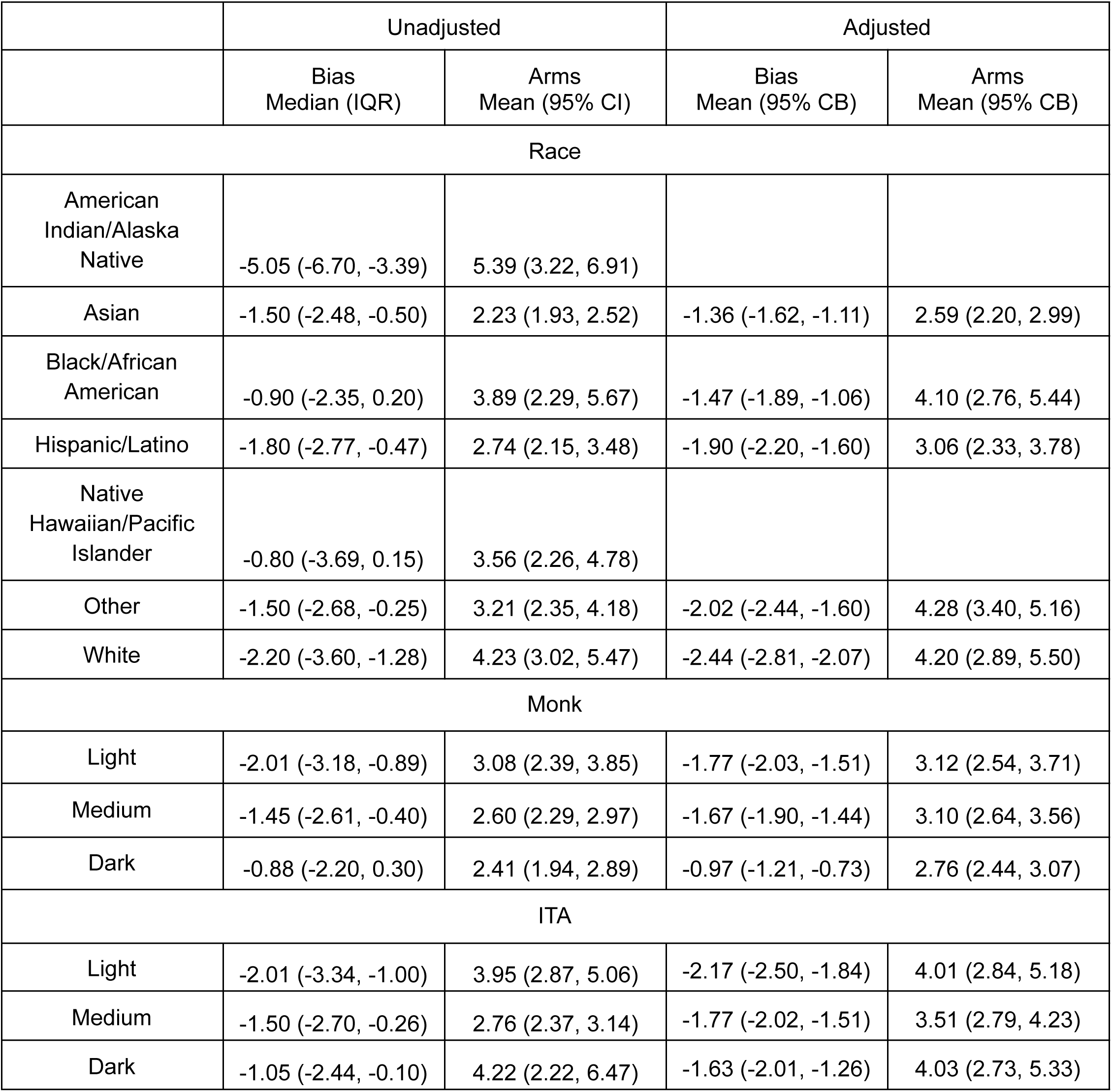
Summary of Bias and ARMS by Group in Full Cohort.

### Association with Skin Pigmentation

The adjusted estimates of bias across skin pigment categories are shown in Figure 3 and Table 2. Adjusted bias associated with the lightest skin pigment groups was significantly more negative than the darkest groups (ITA and MST) (Table E5): a difference 0.54% (95% CB 0.04, 1.04, q-value = 0.02) by ITA and a difference = 0.80%; 95% CB: 0.45, 1.15, q-value <0.001) by MST.

The adjusted estimated A_RMS_ did not differ significantly between skin pigment categories (Figure E4).

### Association with Race and Ethnicity

In the full cohort, the median oximeter bias was negative for all race/ethnicity categories (Table 2, Figure E3). The interquartile range of observed bias included positive bias in participants who identified as Black/African American or Hawaiian/Pacific Islander. The adjusted estimate of mean bias for White patients was −2.44% (95% Confidence Band: - 2.81, −2.07) and significantly more negative than all other race/ethnicity groups except Other (Table 2).

In the full cohort, the adjusted A_RMS_ for all race and ethnicity groups was >3% except among participants identified as Asian. The unadjusted A_RMS_ in the full cohort for Asian and Latino participants groups was <3% (Figure S4, Table 2).

### Subgroup Analyses

The “manufacturer guidance” subgroup (527 patients, 1325 samples) was further divided at the observation level (Figure 1). A patient could contribute data to multiple observation subgroups.

### “Manufacturer Guidance” Subgroup

In the “manufacturer guidance” subgroup pulse oximeter bias was negative, [median - 1.70, IQR (−2.82, −0.50)]. Compared to the full cohort, the A_RMS_ was smaller in the “manufacturer guidance” subgroup, 2.93 (95% CI 2.61, 3.36). Pulse oximeters sensitivity for detecting hypoxemia (SaO_2_ <90%) was 0.69 (95% CI: 0.44, 0.88) and specificity was 0.98 (95% CI: 0.97, 0.99). Overall, adjusted analyses showed similar results between the “manufacturer guidance” and full cohorts (Figures 3, E4 and Tables 2, E6-E9).

### Finger Probe and Ear Probe Subgroups

The magnitude of bias and A_RMS_ were significantly larger for ear compared to finger probes in the full cohort, (−2.20 (IQR −4.00, −0.50) vs. −1.60 (IQR −2.90, −0.20), p-value <0.001) and (4.90 (95% CI 3.90, 6.20) vs. 2.70 (95% CI 2.50, 3.10), p-value <0.001). Similar results comparing ear to finger probes were found in the “manufacturer guidance” subgroup (Figures 3 and E4), and in all skin pigment and race/ethnicity groups (Table E8 and E9, Figure E4).

### Sensitivity Analyses

The relationship between oximeter bias and ITA as a continuous variable was not linear [(Harvey-Collier test, *p < 0.001)*, (rainbow test, p < .001), Figure E5). Analyses with a dark ITA cutoff of <-10° vs <-30° did not significantly change estimates of bias or A_RMS_ for the darkly pigmented grouping. Analyses with a dark ITA cutoff of <-50° vs <-30° found no significant difference between pigment groups. Among patients with ITA <-50° (n=37), we noted a small number of observations with large negative bias. The other sensitivity analyses did not substantially change our results or interpretation.

## Discussion

Here we present findings from the largest prospective cohort study to date of pulse oximeter bias across diverse skin pigments in 631 critically-ill adult patients with directly observed, stable SaO_2_ and SpO_2_ measurements. On average we found negative pulse oximeter bias (SpO_2_ underestimating SaO_2_) across skin pigment groups, with relatively less negative bias in patients with darker pigment. We did not find a linear relationship between bias and skin pigment. Use of objective or subjective pigmentation categorizations showed similar effects on pulse oximeter bias. Stable hypoxemia and “occult hypoxemia” were rare.

Our findings of negative mean bias contrast with most prior studies which reported positive mean bias,^1,12–14,33–39^ often greater in Black patients than White.^1,5,12–14^ A recent prospective pediatric study reported positive bias, greater in children with darker skin.^40^ However, this study used fractional oxygen saturation which could impact the magnitude and direction of the bias. A large retrospective study and recent meta-analysis also estimated positive pooled mean pulse oximeter bias among people with dark skin pigmentation (1.11% [0.29-1.93%]) and Black/African American people (1.52% [0.95-2.09%]).^41,42^ Another review of 33 studies found positive bias of 1.58% for darkly pigmented individuals, nearly double that of individuals with medium or light pigment.^43^

Ours is not the first prospective study to find negative mean bias or smaller bias in Black/African American patients. A 2018 study of 404 critically-ill adults by Ebmeier *et al.* found bias in Masimo pulse oximeter probes of −0.7% (95% CI - 4.8%, 3.3%).^44^ A pediatric study from 2014 reported fewer large absolute pulse oximeter bias readings (>3%) among African American children than White children.^36^ Controlled desaturation laboratory studies in healthy adults suggests that the direction of bias can vary by oximeter brand.^45^ Despite overall negative bias in our data, relative bias between groups was consistent with prior studies. Pulse oximeters read relatively higher for Black/African Americans than Whites and higher in people with dark skin compared to light skin. The significantly higher rate of positive bias in the patients with darker skin pigment is noteworthy. Nearly one-third of readings in the darkest skin pigment groups showed positive bias (29% by ITA, 33% by MST), suggesting that even when mean bias is negative, overestimation of SaO_2_ remains common in these groups.

Several factors likely explain differences between our results and prior studies. We used functional saturation and included only stable readings. Other differences include lack of early-pandemic system strain, use of different oximeter or blood gas devices, and a high median oxygen saturation in our cohort.^46^ The incidence of “occult hypoxemia” depends on the prevalence of SaO_2_ <88% (Figure 2). Our cohort had few stable hypoxemia reading, differing substantially from retrospective studies conducted early in the COVID-19 pandemic,^5,47^ and was more similar to the aforementioned 2018 Ebmeier study in this regard. In the study from Australia and New Zealand by Ebmeier *et al.*, mean SpO_2_ was 95.6% and the authors observed 7 SaO_2_ measurements <89%.^44^

The overall cohort A_RMS_ of 3.87 suggests worse oximeter performance in critically-ill patients compared to healthy adults during controlled laboratory testing used for regulatory clearance. The 2013 FDA guidance recommends A_RMS_ ≤ 3% over SaO_2_ 70-100% for finger probes. Observed A_RMS_ values in lab testing are often well below 3%—particularly at SaO levels above 90%, where most data in our study is concentrated. If the same cutoffs outlined in the new 2025 FDA draft guidance (A_RMS_ 95% CI upper limit <3% in healthy adults) are applied to critically-ill patients, the oximeter performance observed in this study would not be acceptable.^29^ We found the sensitivity for detecting hypoxemia was 72%. Although this is higher than the 43% reported by Ebmeier (43), it is a noteworthy finding that may be lower than most clinicians would expect and was not better in the “Manufacturer’s Guidance” subgroup.

Selecting only observations aligned with manufacturers’ guidance excluded 24% of oximeter readings from the “manufacturer guidance” subgroup (Figure 1, Table E2). Differences in bias and A_RMS_ between the full cohort and the subgroups highlight the need to educate clinicians on potential sources of inaccuracy, especially regarding ear probes, and for manufacturers to identify strategies to better communicate when SpO_2_ readings may be less reliable. Furthermore, these differences identify opportunities for improvement in regulatory guidance approaches and device development.

Strengths of this study include direct observation of proper probe use, simultaneous paired observations, and comprehensive adjustment of potential confounders including vasopressor administration, perfusion index, SaO_2_, and the interaction between these variables. The TMLE procedure combined with Super Learner has robust causal inference and does not rely on the correct specification of a single traditional model like linear regression or generalized estimating equations for validity. We found multiplicity-adjusted, statistically significant differences in bias when comparing light vs dark skin tones (ITA and MST) and Black vs. White race groups. Our study separated race/ethnicity and skin pigment analysis, but did not account for important differences in social or structural factors that may be associated with differences in health or healthcare between race/ethnicity groups.^48–51^

### Study limitations

The major limitations of the study include a low prevalence of stable hypoxemia, limited representation of the darkest range of skin pigmentation (Table 1), and the use of only one oximeter brand. The high median oxygen saturation may mask bias that is only apparent at lower SaO_2_ (Figure E1). While our study enrolled patients with diversity in race/ethnicity categories, the distribution of pigmentation clustered towards the middle of ITA and MST. We analyzed data using previously established ITA cutoffs,^19,52^ which may under-represent diversity in common, darker pigmentation levels.

Another limitation is the inability to account for all factors impacting performance such as signal quality and percent modulation.^53^ Although we adjusted for perfusion index (i.e. ∼percent modulation) and observed a wider spread of bias at very low values (Figure E1), without standardized methods to characterize signal quality and percent modulation across devices, generalizability of our findings is limited.

Several additional factors that limit generalizability, including the single center design and the use of a co-oximeter brand, that differs from those in other studies or clinical settings. ^5,12,33^

## Conclusions

Pulse oximeter bias varies with skin pigmentation. On average we observed negative pulse oximeter bias across all skin pigmentation groups, with less negative bias among patients with darker skin. Pulse oximeters perform worse in critically-ill patients compared with laboratory-controlled verification testing. Pulse oximeter inaccuracy may be larger than clinicians appreciate. Future studies are needed with more stable hypoxemia, more darkly pigmented patients, devices from more manufacturers, and standardized signal quality data collection protocols.

## Supporting information

EquiOx Figures and Supplementary Figures

## Data Availability

All data supporting the findings of this study are openly accessible through the Open Oximetry Project Data Repository.
Data are de-identified and shared under the repository's terms, ensuring adherence to data collection protocols, local IRB approvals, and other relevant regulations. Users can access the data by creating a PhysioNet account and agreeing to the repository's terms of use. For access details, please visit the Open Oximetry website.

https://openoximetry.org/data-repository/

## Acknowledgements

We would additionally like to extend our sincere gratitude to Kumudhini Hendrix, Allison O’Neill, Gene Pennello, Shamsu Hashi, Nick Fong, the Respiratory Care Services and ICU Staff and Nurses at Zuckerberg San Francisco General Hospital and Trauma Center for their dedication and valuable contributions to this project.

## Trial Registration

NCT05554510

## Data Sharing Statement

All data supporting the findings of this study are openly accessible through the Open Oximetry Project Data Repository. Data are de-identified and shared under the repository’s terms, ensuring adherence to data collection protocols, local IRB approvals, and other relevant regulations. Users can access the data by creating a PhysioNet account and agreeing to the repository’s terms of use. For access details, please visit the <u>Open Oximetry website</u>.

---

**Table E1:** Covariates in TMLE Models.

| <u>Propensity Score Covariates</u> | <u>Conditional Bias Estimates</u> |
| --- | --- |
| Age<br>Body Mass Index (BMI)<br>Heart Failure with Reduced Ejection Fraction<br>Diabetes Mellitus<br>Venous Stasis Ulcers<br>End Stage Renal Disease on Hemodialysis<br>Chronic Kidney Disease<br>Anasarca noted by treating clinician<br>Gender<br>Essential Hypertension<br>Pulmonary Hypertension<br>Peripheral Vascular Disease<br>Shock present at enrollment<br>Tobacco Use<br>Valvular Heart Disease classified as Moderate to Severe | All Covariates Included in Propensity Scores<br>SO<sub>2</sub><br>Perfusion Index<br>Administration of Methylene Blue Any Time Prior to Observation<br>Titration of Oxygen in Minutes Prior to ABG Sample Draw<br>Type of Supplemental Oxygen Administration<br>Neuromuscular Blockade Infusion<br>Concurrent Vasopressor Administration<br>Net Fluid Balance<br>PaCO<sub>2</sub><br>pH<br>Hemoglobin concentration<br>Finger Diameter<br>Temperature<br>Probe location (finger vs. ear)<br>Mean Arterial Pressure<br>Plethysmography Quality<br>SpO<sub>2</sub> Stability ≥ 30 Seconds Achieved<br>Quality of Probe Positioning<br>Order of Blood Sample (to account for time)<br>Race (in Skin Pigment Analyses) |

**Table E2.**
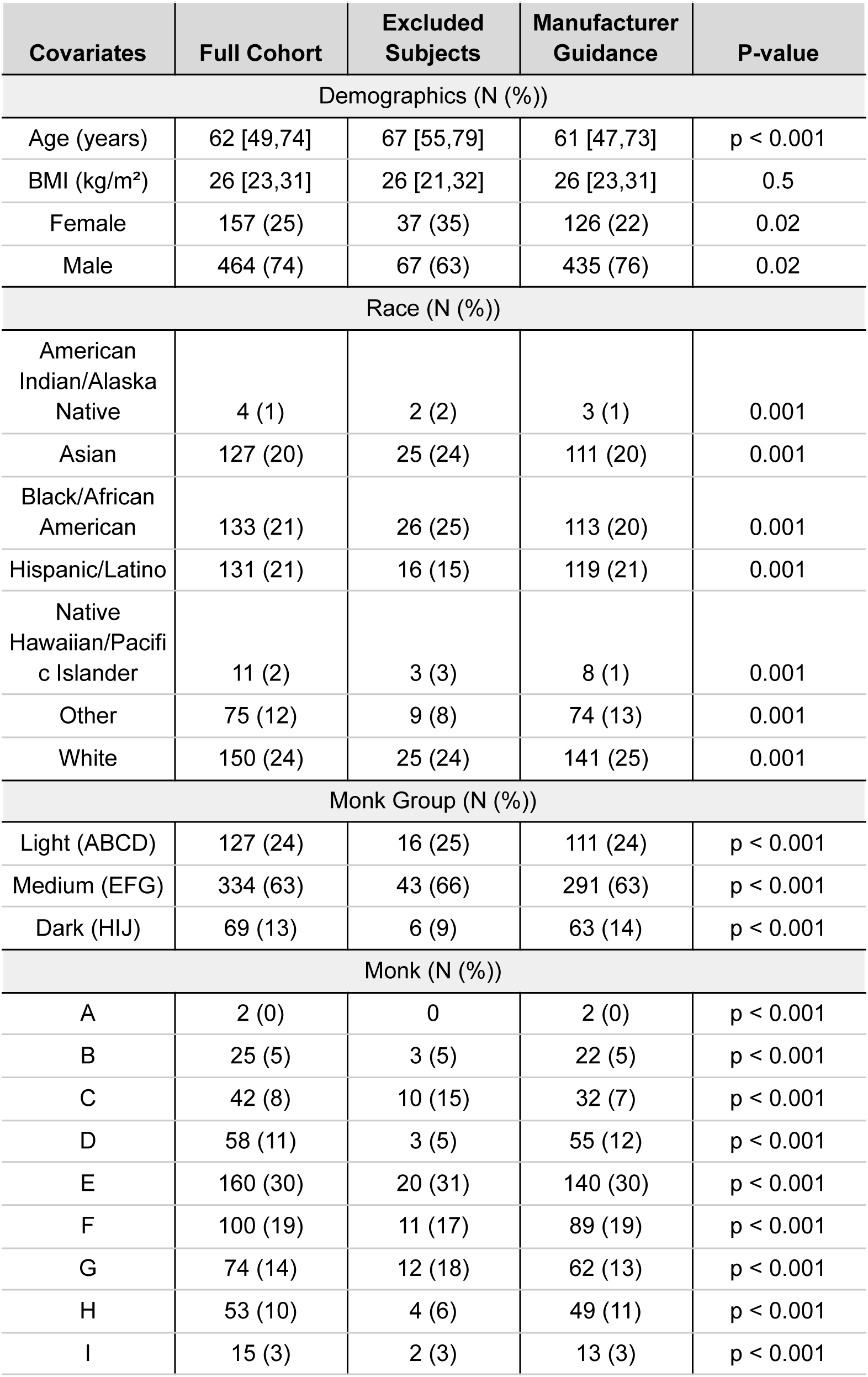
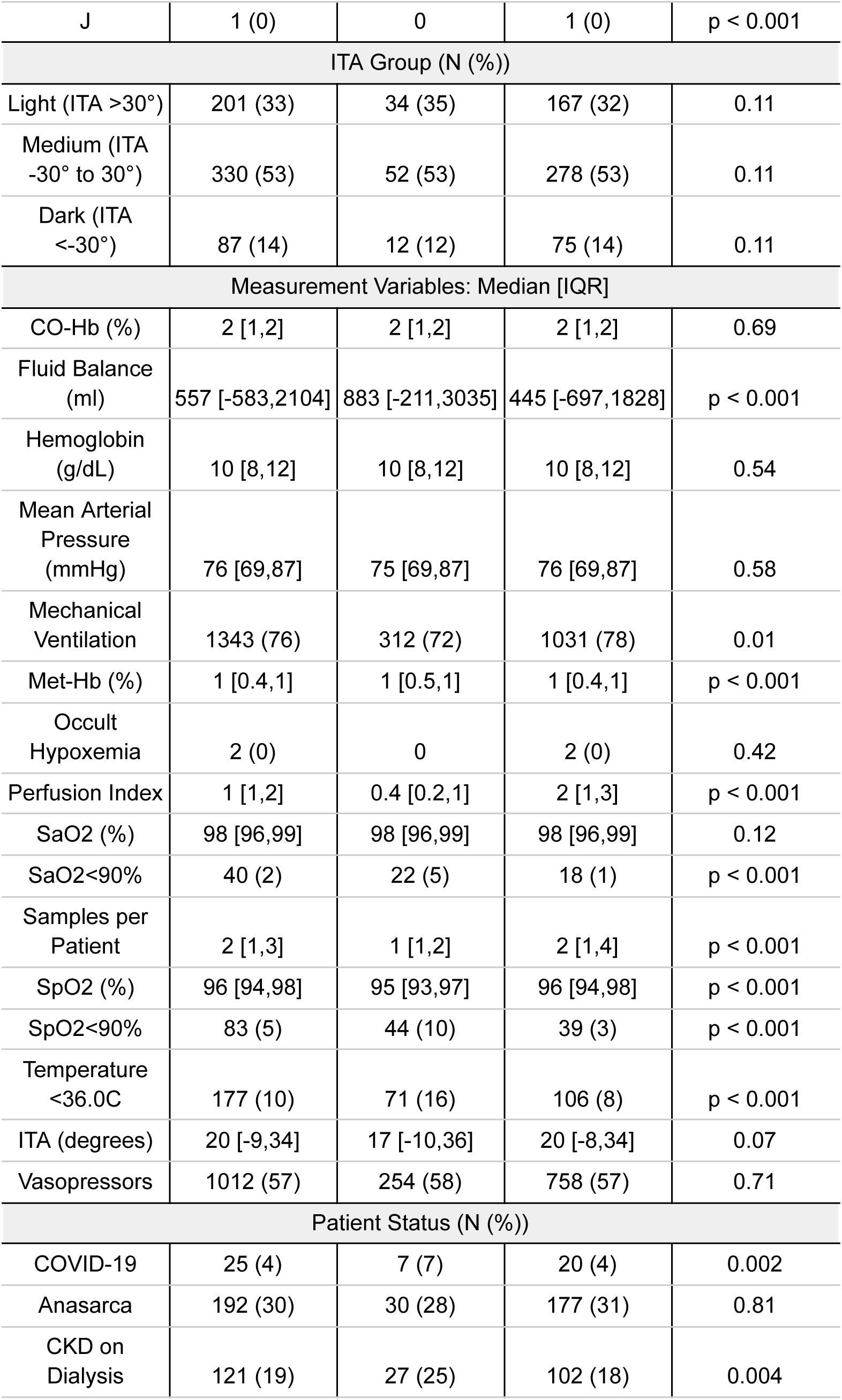
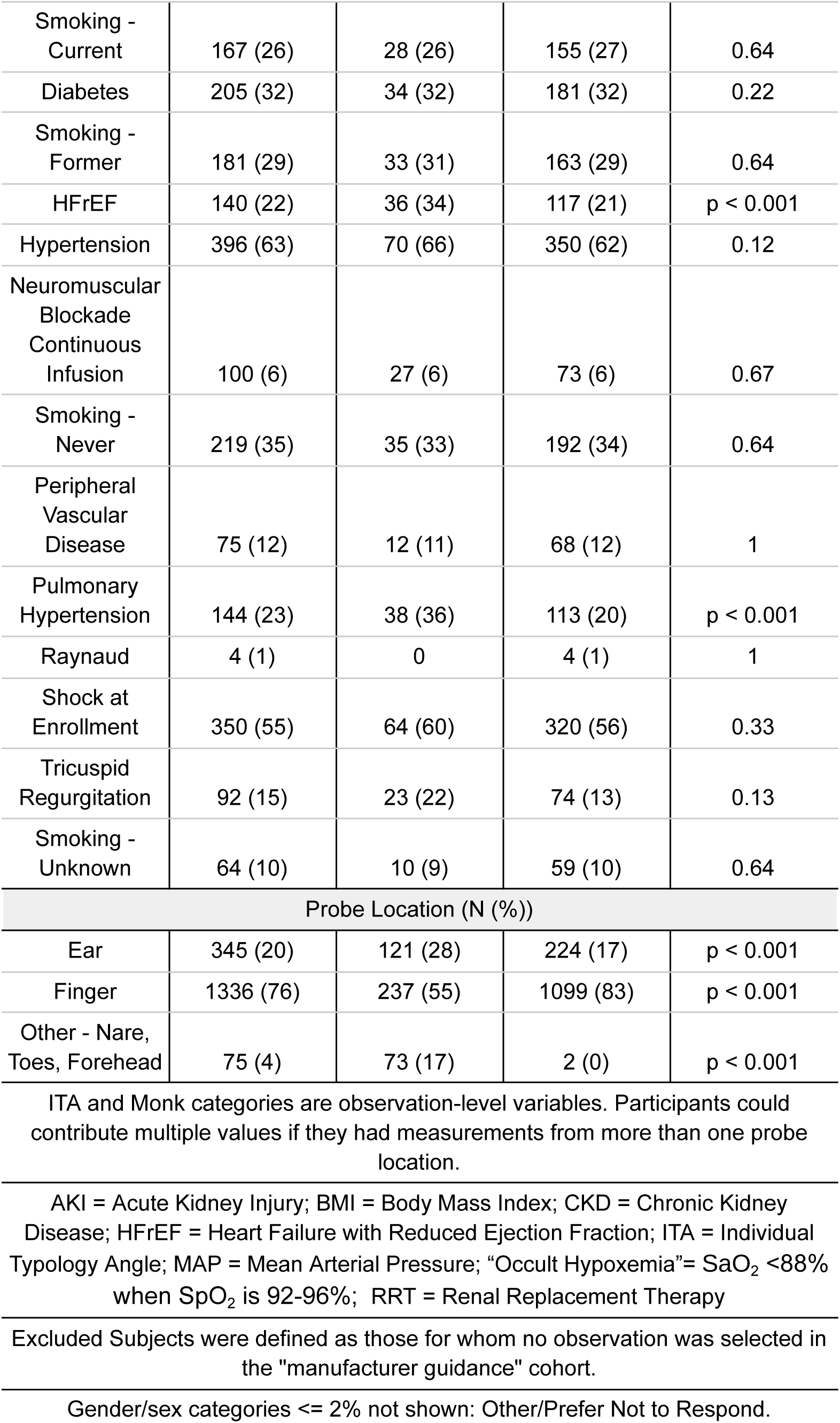
Clinical and Demographic Characteristics of “Manufacturer Guidance” Cohort.

| Covariates | Full Cohort | Excluded Subjects | Manufacturer Guidance | P-value |
| --- | --- | --- | --- | --- |
| Demographics (N (%)) |  |  |  |  |
| Age (years) | 62 [49,74] | 67 [55,79] | 61 [47,73] | p < 0.001 |
| BMI (kg/m<sup>2</sup>) | 26 [23,31] | 26 [21,32] | 26 [23,31] | 0.5 |
| Female | 157 (25) | 37 (35) | 126 (22) | 0.02 |
| Male | 464 (74) | 67 (63) | 435 (76) | 0.02 |
| Race (N (%)) |  |  |  |  |
| American Indian/Alaska Native | 4 (1) | 2 (2) | 3 (1) | 0.001 |
| Asian | 127 (20) | 25 (24) | 111 (20) | 0.001 |
| Black/African American | 133 (21) | 26 (25) | 113 (20) | 0.001 |
| Hispanic/Latino | 131 (21) | 16 (15) | 119 (21) | 0.001 |
| Native Hawaiian/Pacific Islander | 11 (2) | 3 (3) | 8 (1) | 0.001 |
| Other | 75 (12) | 9 (8) | 74 (13) | 0.001 |
| White | 150 (24) | 25 (24) | 141 (25) | 0.001 |
| Monk Group (N (%)) |  |  |  |  |
| Light (ABCD) | 127 (24) | 16 (25) | 111 (24) | p < 0.001 |
| Medium (EFG) | 334 (63) | 43 (66) | 291 (63) | p < 0.001 |
| Dark (HIJ) | 69 (13) | 6 (9) | 63 (14) | p < 0.001 |
| Monk (N (%)) |  |  |  |  |
| A | 2 (0) | 0 | 2 (0) | p < 0.001 |
| B | 25 (5) | 3 (5) | 22 (5) | p < 0.001 |
| C | 42 (8) | 10 (15) | 32 (7) | p < 0.001 |
| D | 58 (11) | 3 (5) | 55 (12) | p < 0.001 |
| E | 160 (30) | 20 (31) | 140 (30) | p < 0.001 |
| F | 100 (19) | 11 (17) | 89 (19) | p < 0.001 |
| G | 74 (14) | 12 (18) | 62 (13) | p < 0.001 |
| H | 53 (10) | 4 (6) | 49 (11) | p < 0.001 |
| I | 15 (3) | 2 (3) | 13 (3) | p < 0.001 |
| J | 1 (0) | 0 | 1 (0) | p < 0.001 |
| ITA Group (N (%)) |  |  |  |  |
| Light (ITA >30°) | 201 (33) | 34 (35) | 167 (32) | 0.11 |
| Medium (ITA -30° to 30°) | 330 (53) | 52 (53) | 278 (53) | 0.11 |
| Dark (ITA <-30°) | 87 (14) | 12 (12) | 75 (14) | 0.11 |
| Measurement Variables: Median [IQR] |  |  |  |  |
| CO-Hb (%) | 2 [1,2] | 2 [1,2] | 2 [1,2] | 0.69 |
| Fluid Balance (ml) | 557 [-583,2104] | 883 [-211,3035] | 445 [-697,1828] | p < 0.001 |
| Hemoglobin (g/dL) | 10 [8,12] | 10 [8,12] | 10 [8,12] | 0.54 |
| Mean Arterial Pressure (mmHg) | 76 [69,87] | 75 [69,87] | 76 [69,87] | 0.58 |
| Mechanical Ventilation | 1343 (76) | 312 (72) | 1031 (78) | 0.01 |
| Met-Hb (%) | 1 [0.4,1] | 1 [0.5,1] | 1 [0.4,1] | p < 0.001 |
| Occult Hypoxemia | 2 (0) | 0 | 2 (0) | 0.42 |
| Perfusion Index | 1 [1,2] | 0.4 [0.2,1] | 2 [1,3] | p < 0.001 |
| SaO2 (%) | 98 [96,99] | 98 [96,99] | 98 [96,99] | 0.12 |
| SaO2<90% | 40 (2) | 22 (5) | 18 (1) | p < 0.001 |
| Samples per Patient | 2 [1,3] | 1 [1,2] | 2 [1,4] | p < 0.001 |
| SpO2 (%) | 96 [94,98] | 95 [93,97] | 96 [94,98] | p < 0.001 |
| SpO2<90% | 83 (5) | 44 (10) | 39 (3) | p < 0.001 |
| Temperature <36.0C | 177 (10) | 71 (16) | 106 (8) | p < 0.001 |
| ITA (degrees) | 20 [-9,34] | 17 [-10,36] | 20 [-8,34] | 0.07 |
| Vasopressors | 1012 (57) | 254 (58) | 758 (57) | 0.71 |
| Patient Status (N (%)) |  |  |  |  |
| COVID-19 | 25 (4) | 7 (7) | 20 (4) | 0.002 |
| Anasarca | 192 (30) | 30 (28) | 177 (31) | 0.81 |
| CKD on Dialysis | 121 (19) | 27 (25) | 102 (18) | 0.004 |
| Smoking - Current | 167 (26) | 28 (26) | 155 (27) | 0.64 |
| Diabetes | 205 (32) | 34 (32) | 181 (32) | 0.22 |
| Smoking - Former | 181 (29) | 33 (31) | 163 (29) | 0.64 |
| HFrEF | 140 (22) | 36 (34) | 117 (21) | p < 0.001 |
| Hypertension | 396 (63) | 70 (66) | 350 (62) | 0.12 |
| Neuromuscular Blockade Continuous Infusion | 100 (6) | 27 (6) | 73 (6) | 0.67 |
| Smoking - Never | 219 (35) | 35 (33) | 192 (34) | 0.64 |
| Peripheral Vascular Disease | 75 (12) | 12 (11) | 68 (12) | 1 |
| Pulmonary Hypertension | 144 (23) | 38 (36) | 113 (20) | p < 0.001 |
| Raynaud | 4 (1) | 0 | 4 (1) | 1 |
| Shock at Enrollment | 350 (55) | 64 (60) | 320 (56) | 0.33 |
| Tricuspid Regurgitation | 92 (15) | 23 (22) | 74 (13) | 0.13 |
| Smoking - Unknown | 64 (10) | 10 (9) | 59 (10) | 0.64 |
| Probe Location (N (%)) |  |  |  |  |
| Ear | 345 (20) | 121 (28) | 224 (17) | p < 0.001 |
| Finger | 1336 (76) | 237 (55) | 1099 (83) | p < 0.001 |
| Other - Nare, Toes, Forehead | 75 (4) | 73 (17) | 2 (0) | p < 0.001 |

ITA and Monk categories are observation-level variables. Participants could contribute multiple values if they had measurements from more than one probe location.
AKI = Acute Kidney Injury; BMI = Body Mass Index; CKD = Chronic Kidney Disease; HFrEF = Heart Failure with Reduced Ejection Fraction; ITA = Individual Typology Angle; MAP = Mean Arterial Pressure; "Occult Hypoxemia" = $\text{SaO}_2 < 88\%$ when $\text{SpO}_2$ is 92-96%; RRT = Renal Replacement Therapy
Excluded Subjects were defined as those for whom no observation was selected in the "manufacturer guidance" cohort.
Gender/sex categories <= 2% not shown: Other/Prefer Not to Respond.

**Table E3.**
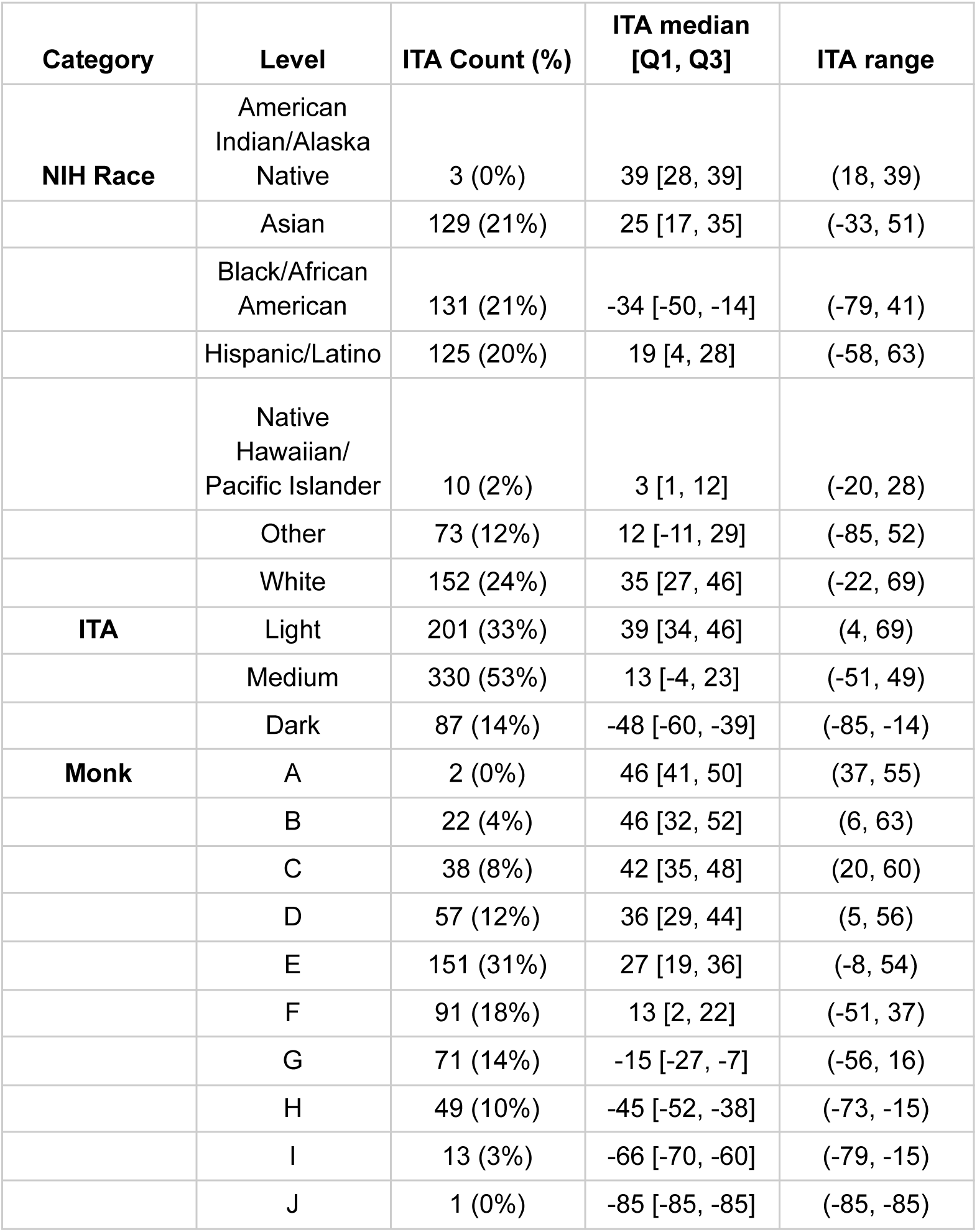
Objective Skin Pigment Data (ITA) by Skin Pigment Categories and Race.

**Table E4.** Clinical and Demographic Characteristics of “Positive Bias” Cohort.

| Covariates | Overall | Non-positive Bias<br>(Bias ≤0) | Positive Bias<br>(Bias >0) | P-value |
| --- | --- | --- | --- | --- |
| Demographics (N (%)) |  |  |  |  |
| Age (years) | 62 [49,74] | 62 [49,75] | 61 [48,72] | 0.04 |
| BMI (kg/m<sup>2</sup>) | 26 [23,31] | 26 [23,31] | 26 [22,31] | 0.94 |
| Female | 157 (25) | 117 (27) | 46 (19) | 0.004 |
| Male | 464 (74) | 311 (71) | 191 (80) | 0.004 |
| Race (N (%)) |  |  |  |  |
| American Indian/Alaska Native | 4 (1) | 5 (1) | 0 | p < 0.001 |
| Asian | 127 (20) | 88 (20) | 48 (20) | p < 0.001 |
| Black/African American | 133 (21) | 70 (16) | 69 (29) | p < 0.001 |
| Hispanic/Latino | 131 (21) | 86 (20) | 49 (21) | p < 0.001 |
| Native Hawaiian/Pacific Islander | 11 (2) | 8 (2) | 3 (1) | p < 0.001 |
| Other | 75 (12) | 53 (12) | 30 (13) | p < 0.001 |
| White | 150 (24) | 126 (29) | 40 (17) | p < 0.001 |
| Monk (N (%)) |  |  |  |  |
| A | 2 (0) | 2 (1) | 0 | p < 0.001 |
| B | 25 (5) | 19 (6) | 6 (3) | p < 0.001 |
| C | 42 (8) | 32 (9) | 10 (5) | p < 0.001 |
| D | 58 (11) | 41 (12) | 17 (9) | p < 0.001 |
| E | 160 (30) | 110 (32) | 50 (26) | p < 0.001 |
| F | 100 (19) | 68 (20) | 32 (17) | p < 0.001 |
| G | 74 (14) | 37 (11) | 37 (19) | p < 0.001 |
| H | 53 (10) | 26 (8) | 27 (14) | p < 0.001 |
| I | 15 (3) | 4 (1) | 11 (6) | p < 0.001 |
| J | 1 (0) | 0 | 1 (1) | p < 0.001 |
| ITA Group (N (%)) |  |  |  |  |
| Light (ITA >30°) | 201 (33) | 150 (38) | 51 (23) | p < 0.001 |
| Medium (ITA<br>-30° to 30°) | 330 (53) | 203 (51) | 127 (58) | p < 0.001 |
| Dark (ITA<br><-30°) | 87 (14) | 45 (11) | 42 (19) | p < 0.001 |
| Measurement Variables: Median [IQR] |  |  |  |  |
| CO-Hb (%) | 2 [1,2] | 2 [1,2] | 1 [1,2] | p < 0.001 |
| Fluid Balance<br>(ml) | 557 [-583,2104] | 581 [-586,2041] | 510 [-556,2211] | 0.72 |
| Hemoglobin<br>(g/dL) | 10 [8,12] | 10 [9,12] | 10 [8,12] | 0.006 |
| Mean Arterial<br>Pressure<br>(mmHg) | 76 [69,87] | 76 [69,86] | 77 [70,91] | 0.04 |
| Mechanical<br>Ventilation | 1343 (76) | 1090 (77) | 253 (72) | 0.04 |
| Met-Hb (%) | 1 [0.4,1] | 1 [0.4,1] | 1 [0.3,1] | 0.03 |
| Occult<br>Hypoxemia | 2 (0) | 0 | 2 (1) | p < 0.001 |
| Perfusion Index | 1 [1,2] | 2 [1,3] | 1 [0.3,2] | p < 0.001 |
| SaO2 (%) | 98 [96,99] | 98 [97,99] | 98 [95,99] | 0.008 |
| SaO2<90% | 40 (2) | 23 (2) | 17 (5) | p < 0.001 |
| Samples per<br>Patient | 2 [1,3] | 2 [1,3] | 3 [2,5] | p < 0.001 |
| SpO2 (%) | 96 [94,98] | 95 [93,97] | 99 [97,100] | p < 0.001 |
| SpO2<90% | 83 (5) | 77 (5) | 6 (2) | 0.005 |
| Temperature<br><36.0C | 177 (10) | 122 (9) | 55 (16) | p < 0.001 |
| ITA (degrees) | 20 [-9,34] | 23 [4,36] | 6 [-22,29] | p < 0.001 |
| Vasopressors | 1012 (57) | 812 (58) | 200 (57) | 0.87 |
| Patient Status (N (%)) |  |  |  |  |
| COVID-19 | 25 (4) | 15 (3) | 12 (5) | 0.42 |
| Anasarca | 192 (30) | 127 (29) | 80 (33) | 0.39 |
| CKD on<br>Dialysis | 121 (19) | 79 (18) | 50 (21) | 0.82 |
| Smoking -<br>Current | 167 (26) | 107 (25) | 76 (32) | p < 0.001 |
| Diabetes | 205 (32) | 112 (26) | 103 (43) | p < 0.001 |
| Smoking - | 181 (29) | 124 (28) | 72 (30) | p < 0.001 |
| Former |  |  |  |  |
| HFrEF | 140 (22) | 95 (22) | 58 (24) | 0.83 |
| Hypertension | 396 (63) | 263 (60) | 157 (66) | 0.008 |
| Neuromuscular<br>Blockade<br>Continuous<br>Infusion | 100 (6) | 81 (6) | 19 (5) | 0.91 |
| Smoking -<br>Never | 219 (35) | 159 (36) | 68 (28) | p < 0.001 |
| Peripheral<br>Vascular<br>Disease | 75 (12) | 41 (9) | 39 (16) | p < 0.001 |
| Pulmonary<br>Hypertension | 144 (23) | 97 (22) | 54 (23) | 0.27 |
| Raynaud | 4 (1) | 2 (0) | 2 (1) | 0.42 |
| Manufacturer<br>Guidance | 1325 (75) | 1075 (76) | 250 (71) | 0.06 |
| Shock at<br>Enrollment | 350 (55) | 233 (53) | 151 (63) | p < 0.001 |
| Tricuspid<br>Regurgitation | 92 (15) | 59 (14) | 38 (16) | 0.35 |
| Smoking -<br>Unknown | 64 (10) | 46 (11) | 23 (10) | p < 0.001 |
| Probe Location (N (%)) |  |  |  |  |
| Ear | 345 (20) | 278 (20) | 67 (19) | 0.03 |
| Finger | 1336 (76) | 1066 (76) | 270 (78) | 0.03 |
| Other - Nare,<br>Toes, Forehead | 75 (4) | 64 (5) | 11 (3) | 0.03 |

ITA and Monk categories are observation-level variables. Participants could contribute multiple values if they had measurements from more than one pulse oximeter probe location.
AKI = Acute Kidney Injury; BMI = Body Mass Index; CKD = Chronic Kidney Disease; HFrEF = Heart Failure with Reduced Ejection Fraction; ITA = Individual Typology Angle; MAP = Mean Arterial Pressure; "Occult Hypoxemia" = $\text{SaO}_2 < 88\%$ when $\text{SpO}_2$ is 92-96% ; RRT = Renal Replacement Therapy
Gender/sex categories <= 2% not shown: Other/Prefer Not to Respond.

**Table E5.** Estimated Differences in Marginal Bias Across Groups in Full Cohort.

| Race | Black / African American | Asian | Hispanic/ Latino | Other |
| --- | --- | --- | --- | --- |
| Asian | -0.11 |  |  |  |
| Hispanic / Latino | 0.43 | **0.54** |  |  |
| Other | **0.55** | **0.66** | 0.12 |  |
| White | **0.97** | **1.08** | **0.54** | 0.42 |
| Monk | Dark | Medium | Light |  |
| Medium | **0.70** |  |  |  |
| Light | **0.80** | 0.10 |  |  |
| ITA | Dark | Medium | Light |  |
| Medium | 0.13 |  |  |  |
| Light | **0.54** | **0.41** |  |  |

**Table E6.** Summary of Bias and ARMS by Group in Manufacturer Guidance Subgroup.

|  | Unadjusted |  | Adjusted |  |
| --- | --- | --- | --- | --- |
|  | Bias<br>Median (IQR) | Arms<br>Mean (95% CI) | Bias<br>Mean (95% CB) | Arms<br>Mean (95% CB) |
| Race |  |  |  |  |
| American<br>Indian/Alaska<br>Native | -3.07 (-3.20, -2.93) | 3.08 (2.80, 3.34) |  |  |
| Asian | -1.50 (-2.28, -0.55) | 2.10 (1.78, 2.45) | -1.52 (-1.75, -1.30) | 2.48 (2.05, 2.90) |
| Black/African<br>American | -0.80 (-2.22, 0.33) | 3.31 (2.10, 4.71) | -1.18 (-1.52, -0.85) | 3.19 (2.28, 4.10) |
| Hispanic/Latino | -1.65 (-2.67, -0.45) | 2.23 (1.99, 2.48) | -1.59 (-1.84, -1.34) | 2.59 (2.28, 2.90) |
| Native<br>Hawaiian/Pacific<br>Islander | -0.55 (-2.16, 0.08) | 2.74 (1.12, 3.82) |  |  |
| Other | -1.90 (-2.69, -0.34) | 2.66 (2.29, 3.11) | -1.75 (-2.07, -1.43) | 2.96 (2.57, 3.36) |
| White | -2.20 (-3.40, -1.31) | 3.02 (2.54, 3.55) | -2.14 (-2.43, -1.86) | 3.17 (2.71, 3.63) |
| Monk |  |  |  |  |
| Light | -2.00 (-2.71, -1.07) | 2.68 (2.20, 3.22) | -1.65 (-1.88, -1.42) | 2.86 (2.45, 3.26) |
| Medium | -1.50 (-2.60, -0.30) | 2.76 (2.20, 3.60) | -1.59 (-1.85, -1.33) | 2.93 (2.28, 3.58) |
| Dark | -0.88 (-1.95, 0.48) | 2.38 (1.90, 2.88) | -1.10 (-1.38, -0.81) | 2.78 (2.32, 3.24) |
| ITA |  |  |  |  |
| Light | -1.95 (-2.90, -1.00) | 2.63 (2.30, 2.96) | -1.81 (-2.04, -1.57) | 2.91 (2.59, 3.22) |
| Medium | -1.56 (-2.53, -0.40) | 2.75 (2.20, 3.53) | -1.67 (-1.95, -1.40) | 2.97 (2.18, 3.75) |
| Dark | -1.00 (-2.50, -0.07) | 3.19 (2.21, 4.36) | -1.40 (-1.69, -1.10) | 2.86 (2.45, 3.28) |

**Table E7.** Summary of Bias and ARMS by Group in SaO2 <99% Cohort.

|  | Unadjusted |  | Adjusted |  |
| --- | --- | --- | --- | --- |
|  | Bias<br>Median (IQR) | Arms<br>Mean (95% CI) | Bias<br>Mean (95% CB) | Arms<br>Mean (95% CB) |
| Race |  |  |  |  |
| American<br>Indian/Alaska<br>Native | -3.23 (-3.45, -3.02) | 3.26 (2.80, 3.67) |  |  |
| Asian | -1.72 (-2.32, -0.77) | 2.25 (1.90, 2.63) | -1.65 (-1.92, -1.38) | 2.55 (2.06, 3.05) |
| Black/African<br>American | -0.75 (-2.28, 0.59) | 3.54 (2.18, 5.14) | -1.00 (-1.36, -0.63) | 3.34 (2.52, 4.16) |
| Hispanic/Latino | -1.57 (-2.72, -0.45) | 2.32 (2.01, 2.70) | -1.53 (-1.82, -1.24) | 2.54 (2.17, 2.91) |
| Native<br>Hawaiian/Pacific<br>Islander | -0.30 (-1.30, 0.15) | 2.08 (0.77, 3.04) |  |  |
| Other | -1.80 (-2.67, -0.22) | 2.81 (2.30, 3.29) | -1.60 (-1.96, -1.23) | 3.02 (2.57, 3.46) |
| White | -2.20 (-3.61, -1.44) | 2.96 (2.56, 3.43) | -2.17 (-2.46, -1.88) | 3.13 (2.72, 3.54) |
| Monk |  |  |  |  |
| Light | -2.10 (-3.40, -1.00) | 2.56 (2.27-2.84) | -1.68 (-1.97, -1.39) | 2.73 (2.40, 3.05) |
| Medium | -1.60 (-2.70, -0.35) | 2.92 (2.30, 3.83) | -1.56 (-1.87, -1.25) | 3.00 (2.08, 3.92) |
| Dark | -0.80 (-1.85, 0.70) | 2.57 (1.96, 3.10) | -0.80 (-1.20, -0.40) | 2.84 (2.07, 3.60) |
| ITA |  |  |  |  |
| Light | -1.95 (-3.00, -1.00) | 2.69 (2.36, 3.05) | -1.83 (-2.09, -1.58) | 2.93 (2.60, 3.25) |
| Medium | -1.60 (-2.58, -0.35) | 2.95 (2.30, 3.84) | -1.67 (-2.02, -1.32) | 3.23 (2.11, 4.35) |
| Dark | -1.20 (-2.42, -0.07) | 3.40 (2.27, 4.71) | -1.20 (-1.53, -0.87) | 2.97 (2.51, 3.43) |

**Table E8.** Summary of Bias and ARMS by Group in Ear Probe Cohort.

|  | Unadjusted |  | Adjusted |  |
| --- | --- | --- | --- | --- |
|  | Bias<br>Median (IQR) | Arms<br>Mean (95% CI) | Bias<br>Mean (95% CB) | Arms<br>Mean (95% CB) |
| Race |  |  |  |  |
| American<br>Indian/Alaska<br>Native | -2.80 (-2.80, -2.80) | 2.80 (2.80, 2.80) |  |  |
| Asian | -1.80 (-2.50, -0.70) | 4.22 (1.71, 6.72) | -2.68 (-3.54, -1.82) | 4.25 (2.81, 5.70) |
| Black/African<br>American | -2.30 (-4.24, 1.07) | 3.94 (3.06, 4.81) | -1.56 (-2.68, -0.44) | 3.89 (3.07, 4.72) |
| Hispanic/Latino | -2.50 (-3.46, -0.30) | 3.06 (2.36, 3.84) | -2.40 (-3.01, -1.79) | 3.77 (3.16, 4.37) |
| Native<br>Hawaiian/Pacific<br>Islander | 0.15 (-0.08, 1.21) | 1.99 (0.15, 3.42) |  |  |
| Other | -2.25 (-3.64, -1.25) | 3.16 (2.12, 4.28) | -1.64 (-2.36, -0.93) | 3.81 (2.94, 4.69) |
| White | -2.60 (-3.82, -1.52) | 4.00 (2.71, 5.26) | -2.28 (-2.93, -1.62) | 3.73 (2.85, 4.62) |
| Monk |  |  |  |  |
| Light | -2.20 (-3.40, -1.38) | 3.52 (2.15, 5.00) | -2.00 (-2.79, -1.21) | 3.57 (2.31, 4.82) |
| Medium | -2.28 (-3.59, -0.40) | 3.17 (2.61, 3.72) | -2.07 (-2.72, -1.41) | 3.56 (2.93, 4.18) |
| Dark | -1.65 (-3.81, 1.41) | 4.28 (2.92, 5.51) | -1.81 (-3.46, -0.16) | 4.33 (3.55, 5.11) |
| ITA |  |  |  |  |
| Light | -2.20 (-3.48, -1.45) | 3.35 (2.40, 4.43) | -2.52 (-3.36, -1.69) | 3.74 (2.87, 4.61) |
| Medium | -2.10 (-3.45, -0.30) | 3.09 (2.59, 3.62) | -1.87 (-2.56, -1.18) | 3.37 (2.78, 3.95) |
| Dark | -3.10 (-5.85, 0.64) | 6.20 (3.57, 8.87) | -3.69 (-5.07, -2.31) | 8.63 (7.38, 9.88) |

**Table E9.** Summary of Bias and ARMS by Group in Finger Probe Cohort.

|  | Unadjusted |  | Adjusted |  |
| --- | --- | --- | --- | --- |
|  | Bias<br>Median (IQR) | Arms<br>Mean (95% CI) | Bias<br>Mean (95% CB) | Arms<br>Mean (95% CB) |
| Race |  |  |  |  |
| American<br>Indian/Alaska<br>Native | -3.34 (-3.34, -3.34) | 3.34 (3.34, 3.34) |  |  |
| Asian | -1.45 (-2.20, -0.50) | 1.98 (1.75, 2.23) | -1.49 (-1.72, -1.25) | 2.28 (1.87, 2.68) |
| Black/African<br>American | -0.75 (-1.80, 0.28) | 3.18 (1.77, 4.76) | -1.29 (-1.63, -0.96) | 2.91 (1.81, 4.00) |
| Hispanic/Latino | -1.60 (-2.62, -0.60) | 2.18 (1.95, 2.40) | -1.49 (-1.72, -1.26) | 2.40 (2.05, 2.75) |
| Native<br>Hawaiian/Pacific<br>Islander | -1.80 (-2.52, -1.30) | 2.19 (0.80, 3.25) |  |  |
| Other | -1.85 (-2.57, -0.26) | 2.53 (2.12, 2.93) | -1.74 (-2.05, -1.43) | 2.70 (2.36, 3.04) |
| White | -2.04 (-3.36, -1.20) | 2.66 (2.37, 2.93) | -1.97 (-2.21, -1.74) | 2.83 (2.52, 3.13) |
| Monk |  |  |  |  |
| Light | -2.00 (-2.72, -1.00) | 2.38 (2.13, 2.63) | -1.49 (-1.70, -1.27) | 2.64 (2.29, 2.99) |
| Medium | -1.45 (-2.40, -0.25) | 2.65 (2.02, 3.47) | -1.50 (-1.75, -1.24) | 2.68 (1.84, 3.52) |
| Dark | -0.78 (-1.77, 0.23) | 1.89 (1.55, 2.19) | -1.06 (-1.29, -0.83) | 2.25 (1.89, 2.61) |
| ITA |  |  |  |  |
| Light | -1.90 (-2.70, -1.00) | 2.42 (2.18, 2.65) | -1.70 (-1.92, -1.48) | 2.69 (2.39, 2.98) |
| Medium | -1.50 (-2.48, -0.50) | 2.73 (2.08, 3.60) | -1.60 (-1.84, -1.35) | 2.74 (1.75, 3.73) |
| Dark | -0.90 (-1.90, -0.07) | 2.09 (1.64, 2.56) | -1.41 (-1.69, -1.12) | 2.52 (1.94, 3.09) |

## References

1. Jubran A, Tobin MJ. Reliability of pulse oximetry in titrating supplemental oxygen therapy in ventilator-dependent patients. Chest. 1990;97(6):1420–1425. doi:10.1378/chest.97.6.1420

2. Bickler PE, Feiner JR, Severinghaus JW. Effects of skin pigmentation on pulse oximeter accuracy at low saturation. Anesthesiology. 2005;102(4):715–719. doi:10.1097/00000542-200504000-00004

3. Feiner JR, Severinghaus JW, Bickler PE. Dark skin decreases the accuracy of pulse oximeters at low oxygen saturation: the effects of oximeter probe type and gender. Anesth Analg. 2007;105(6 Suppl):S18–s23. doi:10.1213/01.ane.0000285988.35174.d9

4. Commissioner O of the. FDA In Brief: FDA warns about limitations and accuracy of pulse oximeters. FDA. Published online February 25, 2021. Accessed March 8, 2025. https://www.fda.gov/news-events/fda-brief/fda-brief-fda-warns-about-limitations-and-accuracy-pulse-oximeters

5. Fawzy A, Wu TD, Wang K, et al. Racial and Ethnic Discrepancy in Pulse Oximetry and Delayed Identification of Treatment Eligibility Among Patients With COVID-19. JAMA Intern Med. 2022;182(7):730–738. doi:10.1001/jamainternmed.2022.1906

6. Gottlieb ER, Ziegler J, Morley K, Rush B, Celi LA. Assessment of Racial and Ethnic Differences in Oxygen Supplementation Among Patients in the Intensive Care Unit. JAMA Intern Med. 2022;182(8):849–858. doi:10.1001/jamainternmed.2022.2587

7. Sudat SEK, Wesson P, Rhoads KF, et al. Racial Disparities in Pulse Oximeter Device Inaccuracy and Estimated Clinical Impact on COVID-19 Treatment Course. Am J Epidemiol. 2023;192(5):703–713. doi:10.1093/aje/kwac164

8. Seitz KP, Wang L, Casey JD, et al. Pulse Oximetry and Race in Critically Ill Adults. Crit Care Explor. 2022;4(9):e0758. doi:10.1097/CCE.0000000000000758

9. ISO, IEC. Medical Electrical Equipment: Particular Requirements for Basic Safety and Essential Performance of Pulse Oximeter Equipment.; 2017. https://www.iso.org/obp/ui/#iso:std:iso:80601:-2-61:ed-2:v2:en

10. Administration USF and D. Pulse Oximeters - Premarket Notification Submissions [510(k)s]: Guidance for Industry and Food and Drug Administration Staff. 2013. https://www.fda.gov/regulatory-information/search-fda-guidance-documents/pulse-oximeters-premarket-notification-submissions-510ks-guidance-industry-and-food-and-drug

11. Sjoding MW, Iwashyna TJ, Valley TS. Change the Framework for Pulse Oximeter Regulation to Ensure Clinicians Can Give Patients the Oxygen They Need. Am J Respir Crit Care Med. 2023;207(6):661–664. doi:10.1164/rccm.202209-1773ED

12. Sjoding MW, Dickson RP, Iwashyna TJ, Gay SE, Valley TS. Racial Bias in Pulse Oximetry Measurement. N Engl J Med. 2020;383(25):2477–2478. doi:10.1056/NEJMc2029240

13. Valbuena VSM, Barbaro RP, Claar D, et al. Racial Bias in Pulse Oximetry Measurement Among Patients About to Undergo Extracorporeal Membrane Oxygenation in 2019-2020: A Retrospective Cohort Study. Chest. 2022;161(4):971–978. doi:10.1016/j.chest.2021.09.025

14. Valbuena VSM, Seelye S, Sjoding MW, et al. Racial bias and reproducibility in pulse oximetry among medical and surgical inpatients in general care in the Veterans Health Administration 2013-19: multicenter, retrospective cohort study. BMJ. 2022;378:e069775. doi:10.1136/bmj-2021-069775

15. Chesley CF, Lane-Fall MB, Panchanadam V, et al. Racial disparities in occult hypoxemia and clinically based mitigation strategies to apply in advance of technological advancements. Respir Care. Published online June 3, 2022. doi:10.4187/respcare.09769

16. Pennello G. In Real-World Studies, Comparisons of Predictive Values Are Confounded by Prevalence. CHEST. 2022;162(2):e103. doi:10.1016/j.chest.2022.03.054

17. Vasudevan S, Vogt WC, Weininger S, Pfefer TJ. Melanometry for objective evaluation of skin pigmentation in pulse oximetry studies. Commun Med. 2024;4(1):138. doi:10.1038/s43856-024-00550-7

18. Study Details | Evaluating Pulse Oximetry Bias in Children With Darker Skin Pigmentation | ClinicalTrials.gov. Accessed January 31, 2025. https://clinicaltrials.gov/study/NCT05617547

19. Del Bino S. Variations in skin colour and the biological consequences of ultraviolet radiation exposure. Br J Dermatol. 2013;169:33–40.

20. Oximetry O. Study Protocols. 2024. https://openoximetry.org/study-protocols/

21. Monk E. The Monk Skin Tone Scale. Published online May 4, 2023. doi:10.31235/osf.io/pdf4c

22. Lipnick MS, Chen D, Law T, et al. Comparison of methods for characterizing skin pigment diversity in research cohorts. Published online February 25, 2025. doi:10.1101/2025.02.21.25322707

23. Committee on the Use of Race and Ethnicity in Biomedical Research, Board on Health Sciences Policy, Board on Population Health and Public Health Practice, Board on Health Care Services, Health and Medicine Division, National Academies of Sciences, Engineering, and Medicine. Rethinking Race and Ethnicity in Biomedical Research. (Wilson MR, Beachy SH, Schumm SN, eds.). National Academies Press; 2025:27913. doi:10.17226/27913

24. Powe NR, Yearby R, Wilson MR. Race and Ethnicity in Biomedical Research: Changing Course and Improving Accountability. JAMA. Published online February 10, 2025. doi:10.1001/jama.2024.28390

25. Revisions to OMB’s Statistical Policy Directive No. 15: Standards for Maintaining, Collecting, and Presenting Federal Data on Race and Ethnicity. Federal Register. March 29, 2024. Accessed February 15, 2025. https://www.federalregister.gov/documents/2024/03/29/2024-06469/revisions-to-ombs-statistical-policy-directive-no-15-standards-for-maintaining-collecting-and

26. NIH Staff. Federal Race and Ethnicity Categories Have Changed – What’s Next? – NIH Extramural Nexus. December 19, 2024. Accessed February 22, 2025. https://nexus.od.nih.gov/all/2024/12/19/federal-race-and-ethnicity-categories-have-changed-whats-next/

27. Harris PA, Taylor R, Minor BL, et al. The REDCap consortium: Building an international community of software platform partners. J Biomed Inform. 2019;95:103208. doi:10.1016/j.jbi.2019.103208

28. Harris PA, Taylor R, Thielke R, Payne J, Gonzalez N, Conde JG. Research electronic data capture (REDCap)--a metadata-driven methodology and workflow process for providing translational research informatics support. J Biomed Inform. 2009;42(2):377–381. doi:10.1016/j.jbi.2008.08.010

29. U.S. Food and Drug Administration. Pulse Oximeters for Medical Purposes - Non-Clinical and Clinical Performance Testing, Labeling, and Premarket Submission Recommendations. US Food Drug Adm. Published online January 7, 2025. Accessed February 15, 2025. https://www.fda.gov/media/184896/download

30. Bland JM, Altman DG. Comparing methods of measurement: why plotting difference against standard method is misleading. Lancet. 1995;346(8982):1085–1087. doi:10.1016/s0140-6736(95)91748-9

31. Balzer LB, Zheng W, van der Laan MJ, Petersen ML. A new approach to hierarchical data analysis: Targeted maximum likelihood estimation for the causal effect of a cluster-level exposure. Stat Methods Med Res. 2019;28(6):1761–1780. doi:10.1177/0962280218774936

32. Van der Laan MJ, Rose S. Targeted Learning: Causal Inference for Observational and Experimental Data. Springer New York; 2011. 10.1007/978-1-4419-9782-1

33. Henry NR, Hanson AC, Schulte PJ, et al. Disparities in Hypoxemia Detection by Pulse Oximetry Across Self-Identified Racial Groups and Associations With Clinical Outcomes. Crit Care Med. 2022;50(2):204–211. doi:10.1097/CCM.0000000000005394

34. Fawzy A, Ali H, Dziedzic PH, et al. Skin Pigmentation and Pulse Oximeter Accuracy in the Intensive Care Unit: a Pilot Prospective Study. medRxiv. Published online November 17, 2023. doi:10.1101/2023.11.16.23298645

35. Louw A, Cracco C, Cerf C, et al. Accuracy of pulse oximetry in the intensive care unit. Intensive Care Med. 2001;27(10):1606–1613. doi:10.1007/s001340101064

36. Ross PA, Newth CJL, Khemani RG. Accuracy of Pulse Oximetry in Children. Pediatrics. 2014;133(1):22–29. doi:10.1542/peds.2013-1760

37. Hao S, Dempsey K, Matos J, et al. Utility of Skin Tone on Pulse Oximetry in Critically Ill Patients: A Prospective Cohort Study. Crit Care Explor. 2024;6(9):e1133. doi:10.1097/CCE.0000000000001133

38. Gd P, Df M, S G, H R, F G. Do changes in pulse oximeter oxygen saturation predict equivalent changes in arterial oxygen saturation? Crit Care Lond Engl. 2003;7(4). doi:10.1186/cc2339

39. Ak S, Ms S, B M, S R. Comparative Evaluation of Accuracy of Pulse Oximeters and Factors Affecting Their Performance in a Tertiary Intensive Care Unit. J Clin Diagn Res JCDR. 2017;11(6). doi:10.7860/JCDR/2017/24640.9961

40. Starnes JR, Welch W, Henderson CC, et al. Pulse Oximetry and Skin Tone in Children. N Engl J Med. 0(0). doi:10.1056/NEJMc2414937

41. Wong AI, Charpignon M, Kim H, et al. Analysis of Discrepancies Between Pulse Oximetry and Arterial Oxygen Saturation Measurements by Race and Ethnicity and Association With Organ Dysfunction and Mortality. JAMA Netw Open. 2021;4(11):e2131674. doi:10.1001/jamanetworkopen.2021.31674

42. Shi C, Goodall M, Dumville J, et al. The accuracy of pulse oximetry in measuring oxygen saturation by levels of skin pigmentation: a systematic review and meta-analysis. BMC Med. 2022;20(1):267. doi:10.1186/s12916-022-02452-8

43. Graham HR, King C, Rahman AE, et al. Reducing global inequities in medical oxygen access: the Lancet Global Health Commission on medical oxygen security. Lancet Glob Health. 2025;0(0). doi:10.1016/S2214-109X(24)00496-0

44. Ebmeier SJ, Barker M, Bacon M, et al. A two centre observational study of simultaneous pulse oximetry and arterial oxygen saturation recordings in intensive care unit patients. Anaesth Intensive Care. 2018;46(3):297–303. doi:10.1177/0310057X1804600307

45. openoximetry.org. Oximeters. OpenOximetry. Accessed February 13, 2025. https://openoximetry.org/oximeters/

46. Weininger S. Effective Standards and Regulatory Tools for Respiratory Gas Monitors and Pulse Oximeters: The Role of the Engineer and Clinician. Anesth Analg. 2007;105(6):S95–S99. doi:10.1213/01.ane.0000278133.48241.0b

47. Sjoding MW, Dickson RP, Iwashyna TJ, Gay SE, Valley TS. Racial Bias in Pulse Oximetry Measurement. N Engl J Med. 2020;383(25):2477–2478. doi:10.1056/NEJMc2029240

48. Monk EP. The Cost of Color: Skin Color, Discrimination, and Health among African-Americans. Am J Sociol. 2015;121(2):396–444. doi:10.1086/682162

49. Eneanya ND, Yang W, Reese PP. Reconsidering the Consequences of Using Race to Estimate Kidney Function. JAMA. 2019;322(2):113. doi:10.1001/jama.2019.5774

50. Bhakta NR, Bime C, Kaminsky DA, et al. Race and Ethnicity in Pulmonary Function Test Interpretation: An Official American Thoracic Society Statement. Am J Respir Crit Care Med. 2023;207(8):978–995. doi:10.1164/rccm.202302-0310ST

51. Care I of M (US) C on U and ER and ED in H, Smedley BD, Stith AY, Nelson AR. Abstract. In: Unequal Treatment: Confronting Racial and Ethnic Disparities in Health Care. National Academies Press (US); 2003. Accessed February 22, 2025. https://www.ncbi.nlm.nih.gov/books/NBK220366/

52. Chardon A, Cretois I, Hourseau C. Skin colour typology and suntanning pathways. Int J Cosmet Sci. 1991;13(4):191–208. doi:10.1111/j.1467-2494.1991.tb00561.x

53. Gudelunas MK, Lipnick M, Hendrickson C, et al. Low Perfusion and Missed Diagnosis of Hypoxemia by Pulse Oximetry in Darkly Pigmented Skin: A Prospective Study. Anesth Analg. 2024;138(3):552–561. doi:10.1213/ANE.0000000000006755

