## Supplementary material for "EquiOx: A Prospective study of pulse oximeter bias and skin pigmentation in critically-ill adults": EquiOx Figures and Supplementary Figures

Figure 1. Selection of “Reasonable Clinician” Cohort for Subgroup Analysis

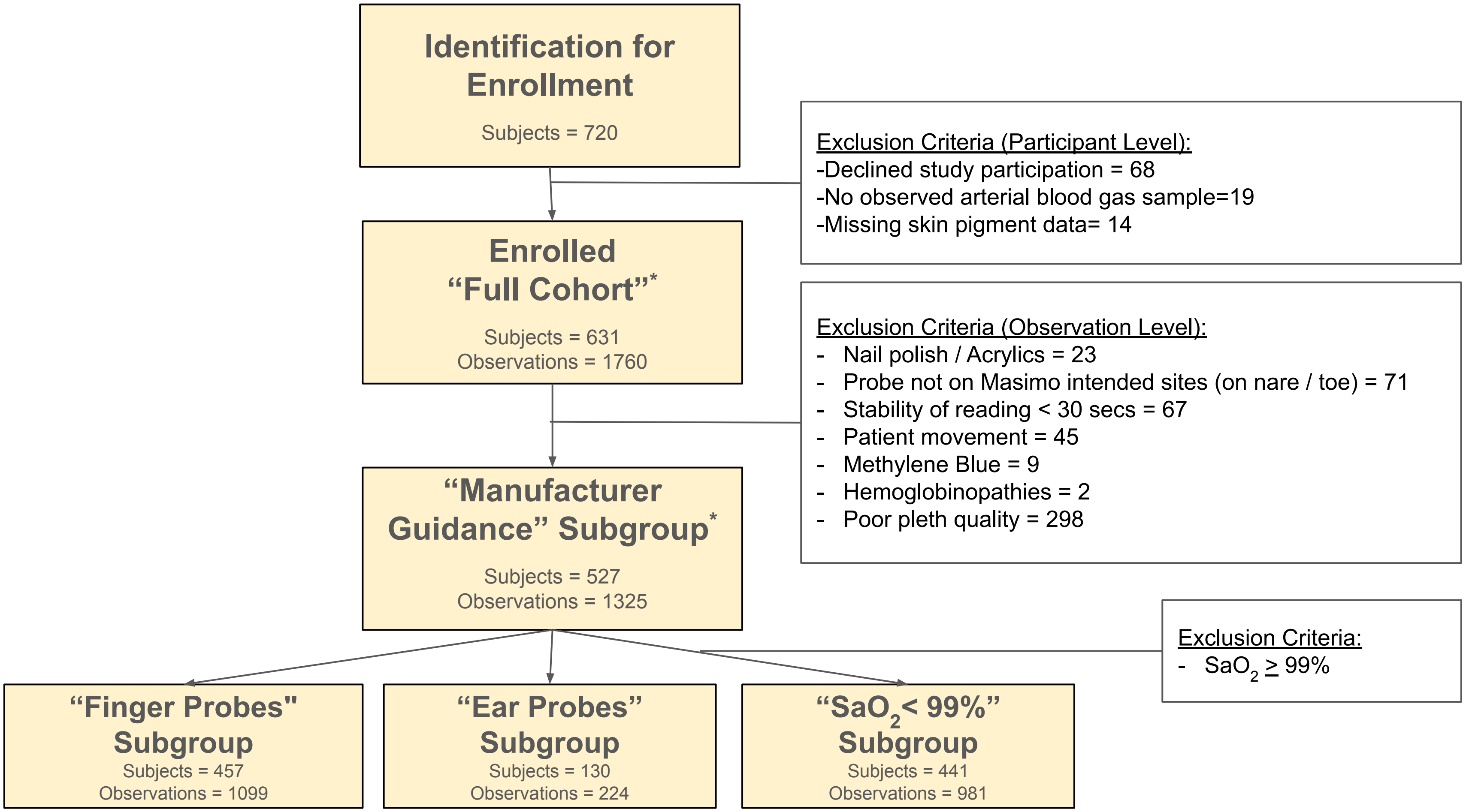

Figure 2. $SaO_2$ Distribution by $SpO_2$ Value Across Race and Ethnicity and Skin Pigment Categories.

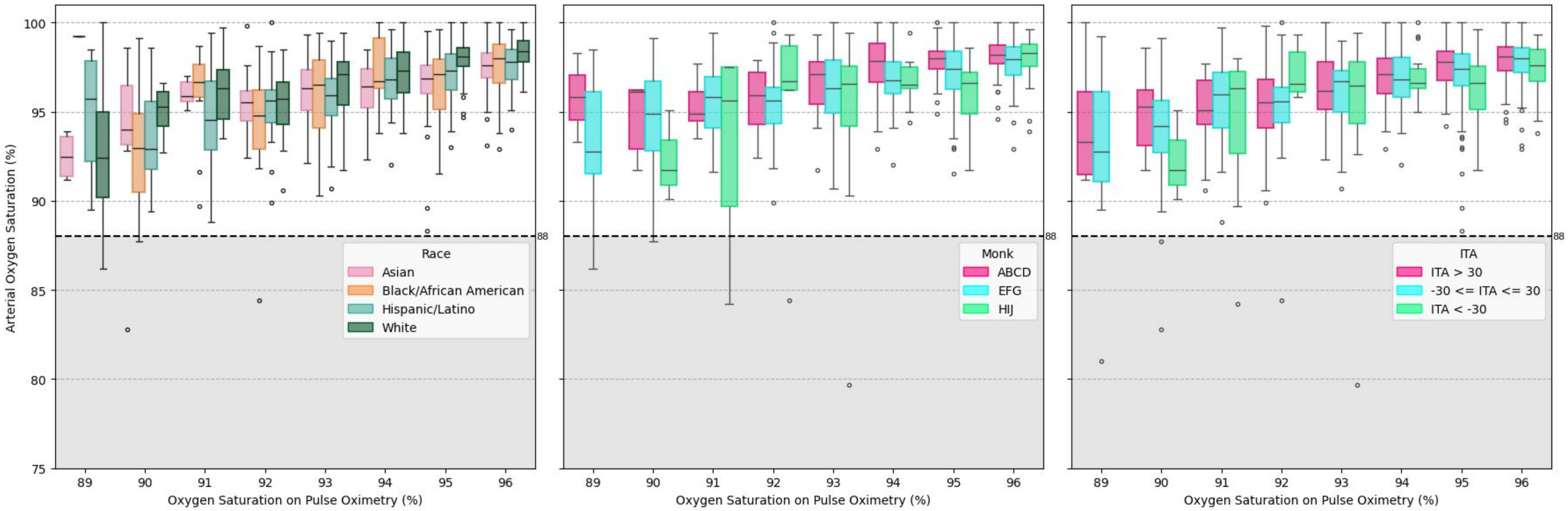

Figure 3. TMLE Adjusted Estimates of Pulse Oximeter Bias by Skin Pigment and Race and Ethnicity.

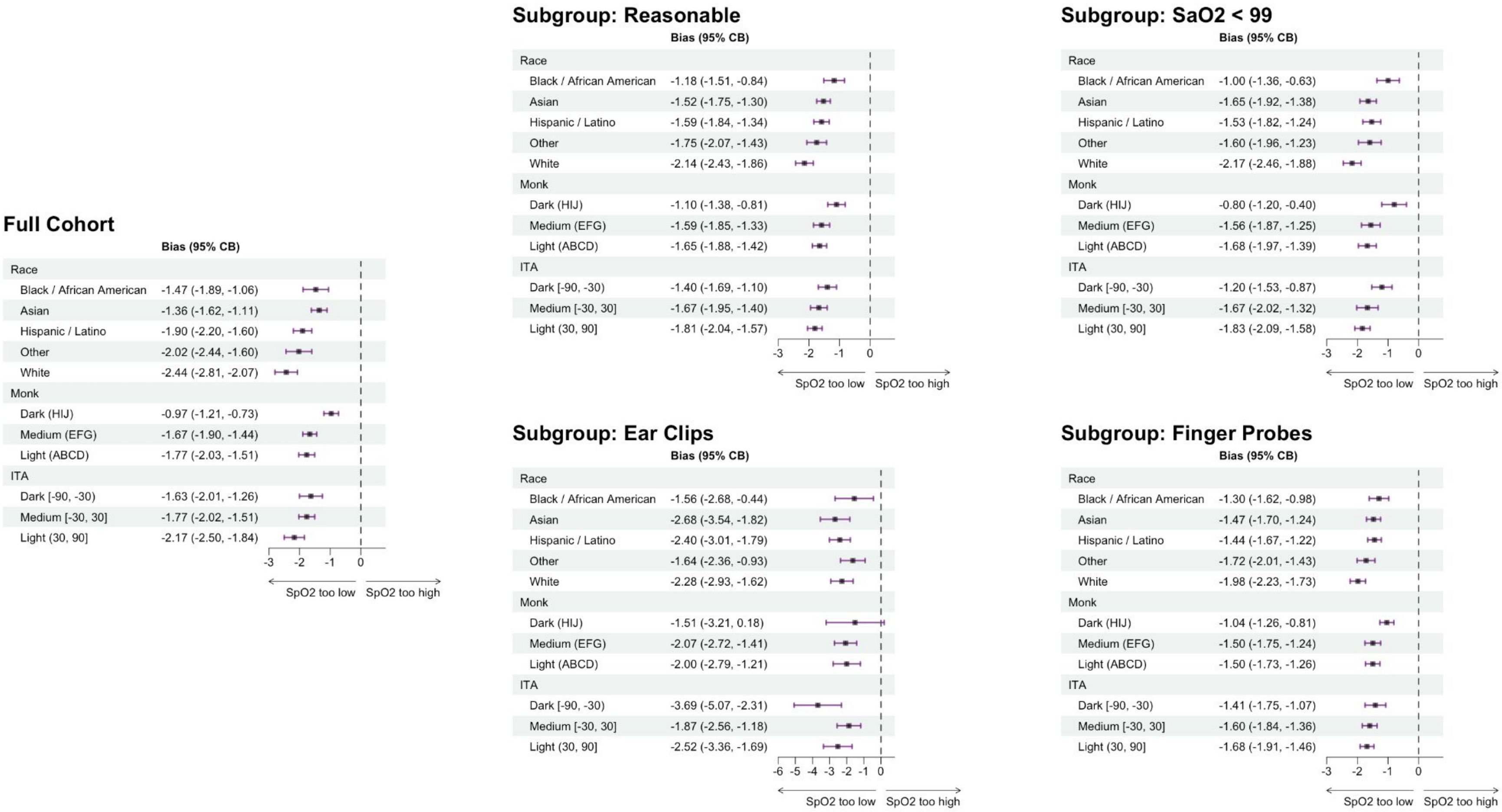

### Supplemental Tables and Figures

Figure E1. Raw Data Distribution of Bias by Skin Pigment and Perfusion Index.

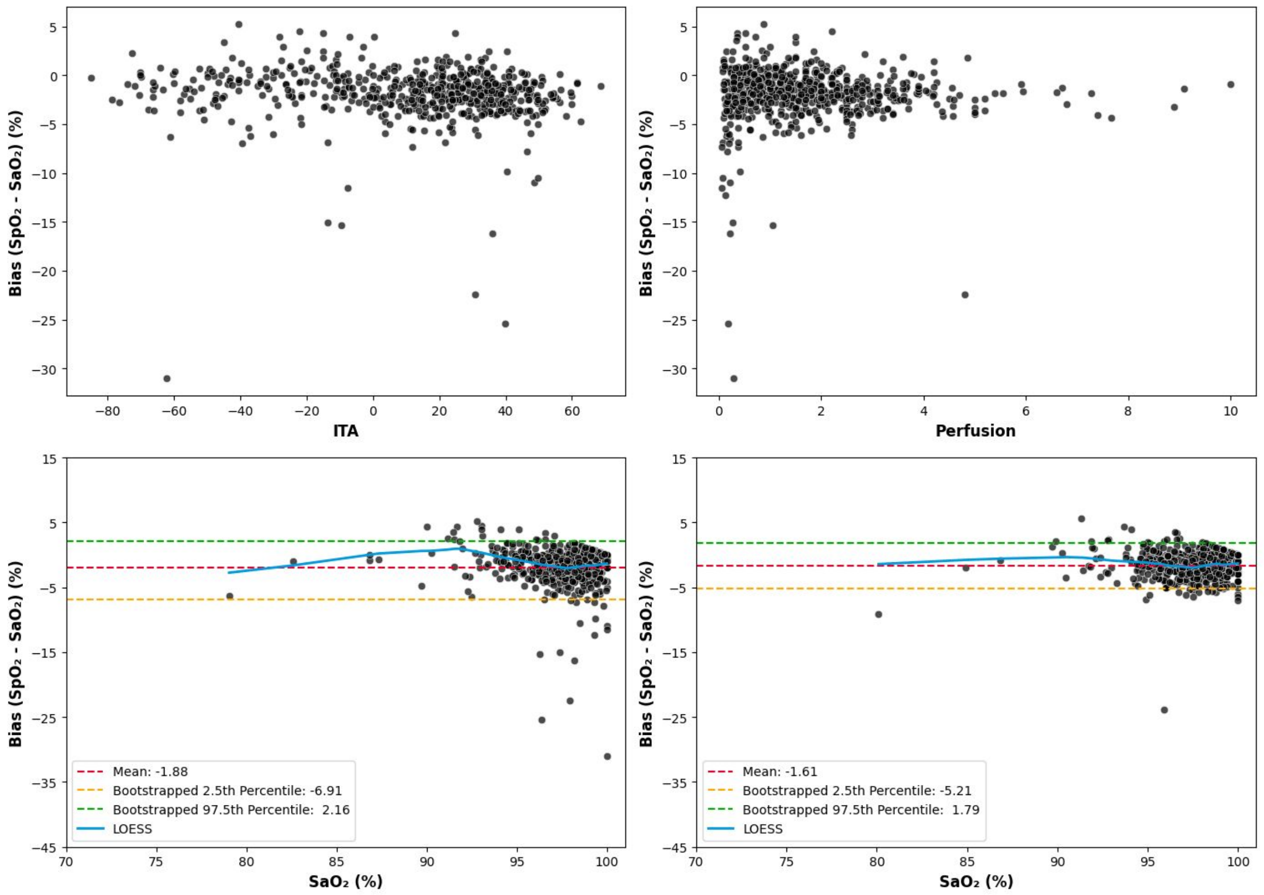

Figure E2. Distribution of $SpO_2$ Bias by Skin Pigment and Race and Ethnicity in Full Cohort.

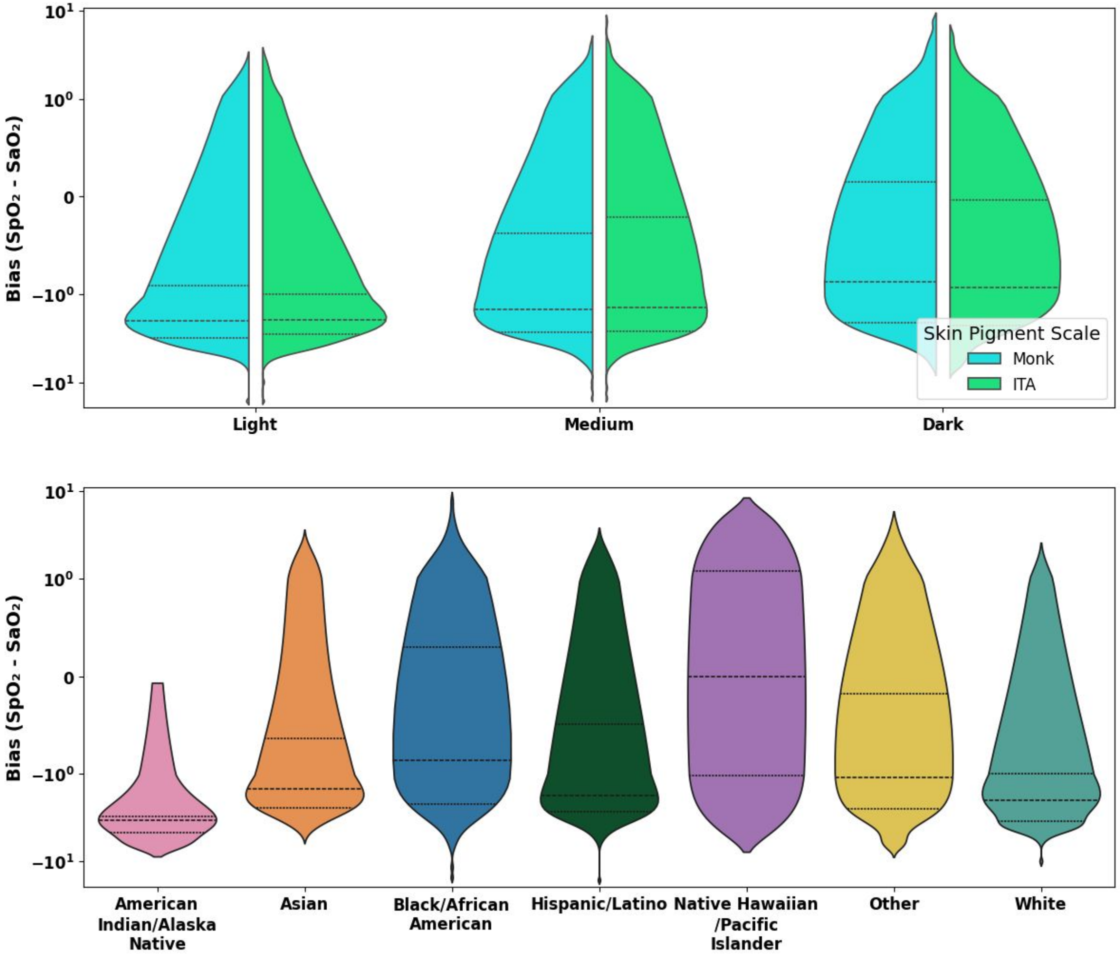

Figure E3. Observed Pulse Oximeter Bias by Skin Pigment and Race and Ethnicity.

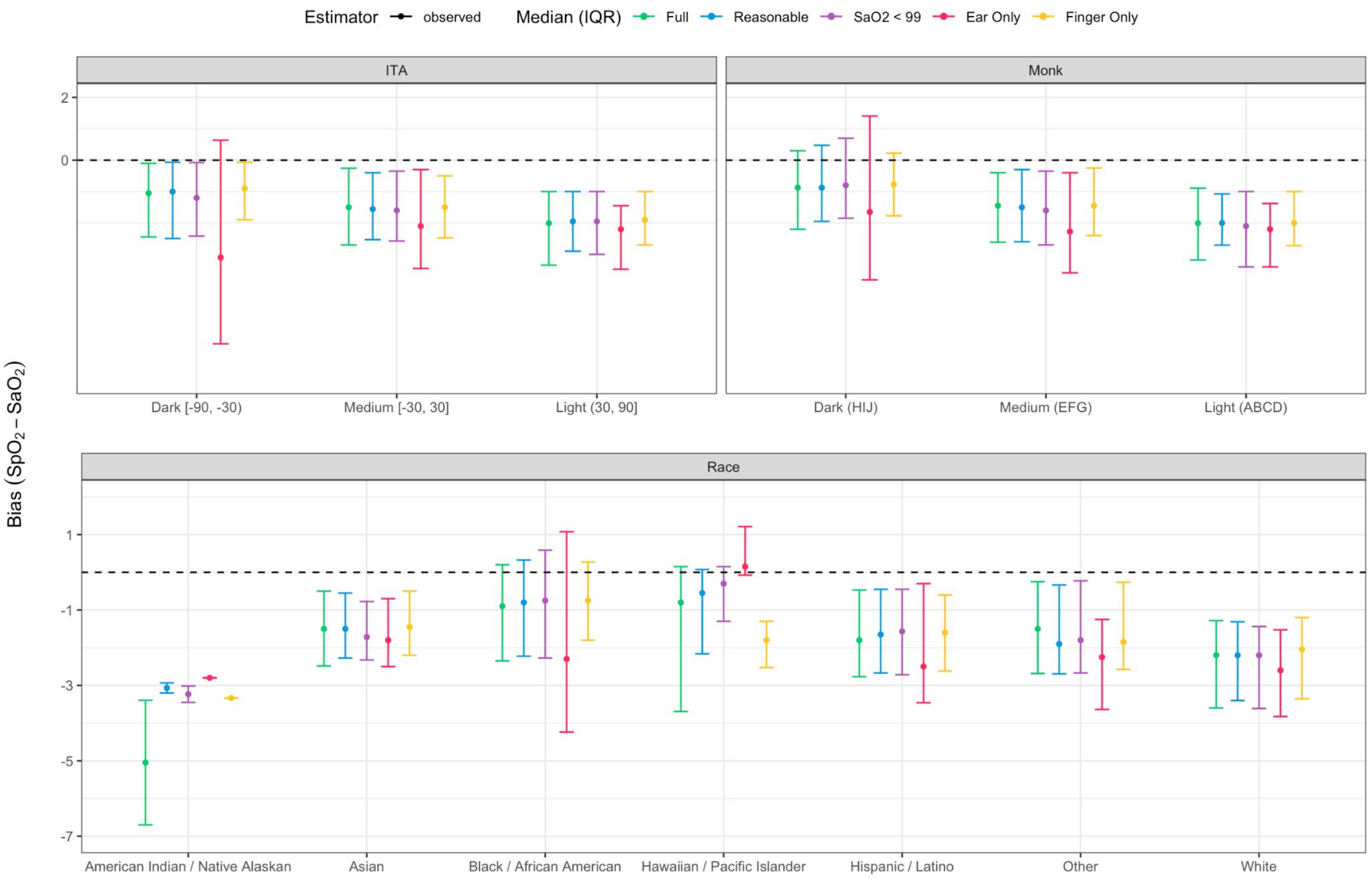

### Figure E4. TMLE-Adjusted Estimates of $A_{RMS}$ by Skin Pigment and Race and Ethnicity.

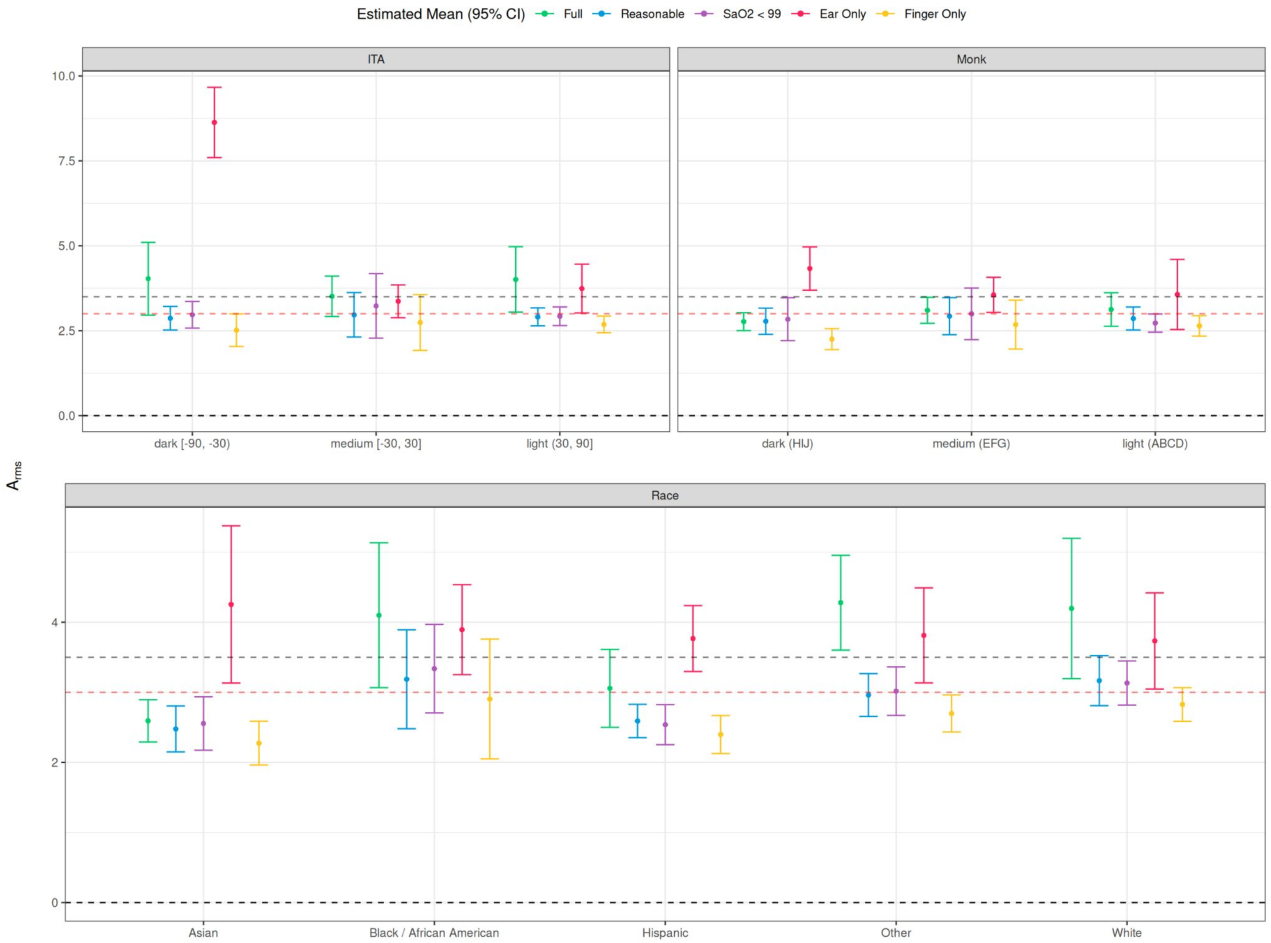

Figure E5. $SpO_2$ Bias vs. ITA

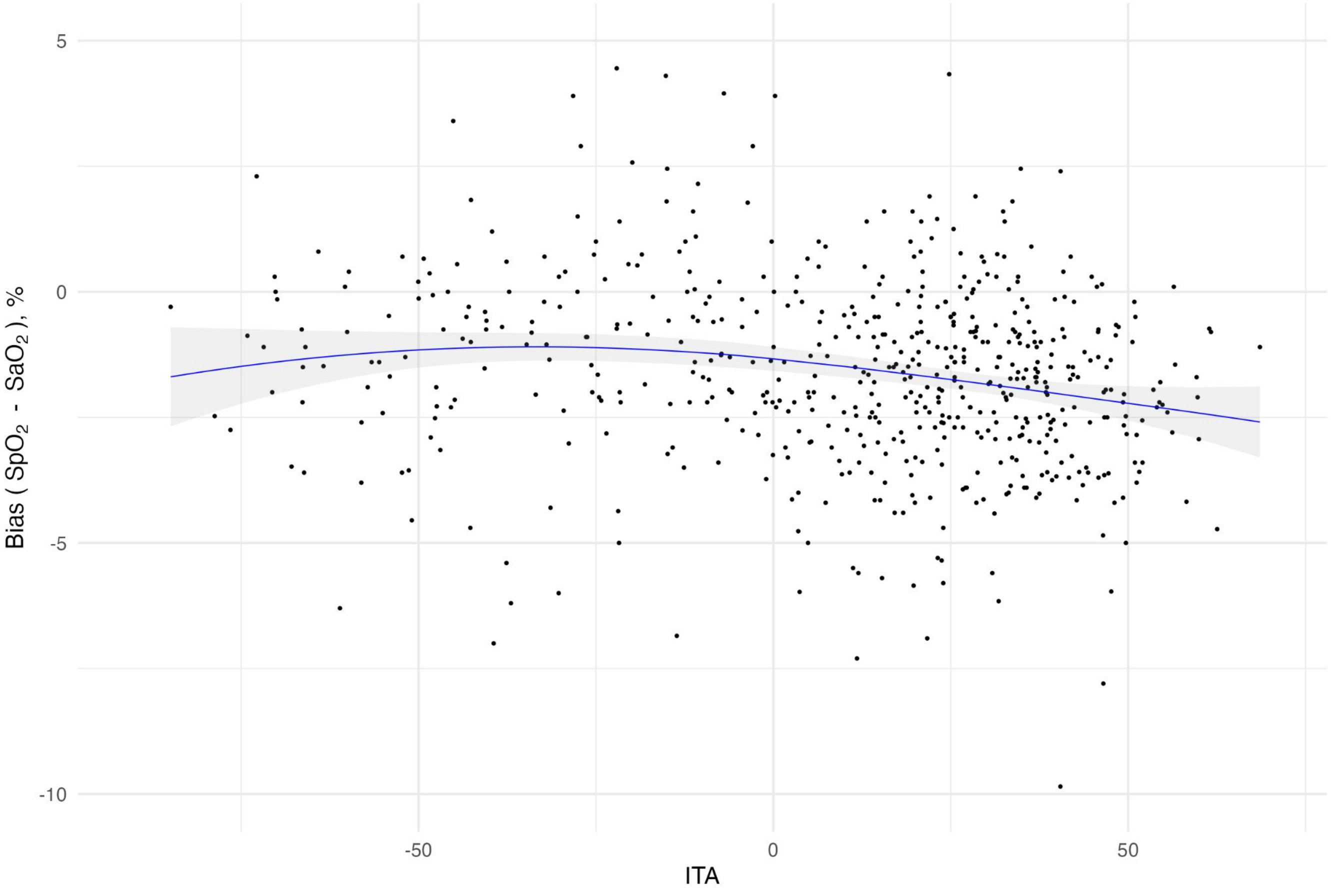

### Figure E6: TMLE Adjusted estimates of pulse oximeter bias by skin pigment and race

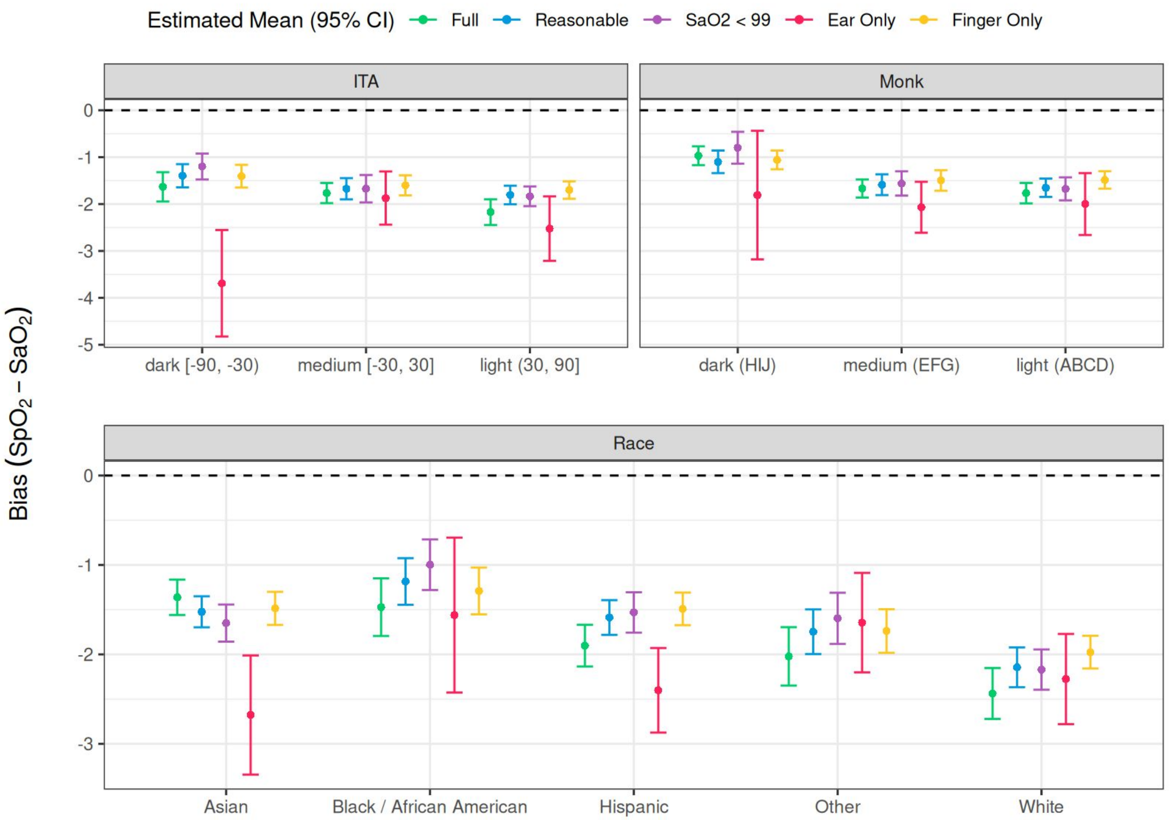
